# Diversification without convergence: national childhood respiratory pathogen spectra diverge as they diversify, 1990–2023

**DOI:** 10.64898/2026.09.01.26361890

**Authors:** Deze Li, Hao Chen, Xiaotong Wang, Chen Shen

## Abstract

**Background:** National childhood respiratory pathogen spectra are diversifying nearly everywhere — within-country diversity rose in 203 of 204 countries between 1990 and 2023 — yet whether countries are diversifying toward a common spectrum or along divergent paths is unknown. We quantified between-country compositional distance of national pathogen spectra over the same period.

**Methods:** We built national pathogen share vectors from Global Burden of Disease Study 2023 lower respiratory infection etiologic attributions (26 pathogens, 204 countries, ages 0–19 years) at five timepoints spanning 1990–2023. Between-country distance was measured as all pairwise Jensen–Shannon divergences (JSD; primary) and Bray–Curtis dissimilarities, with Baselga and Jaccard decompositions; robustness was assessed across metrics, pathogen panels, low-count thresholds and a balanced panel of 107 countries.

**Results:** Mean pairwise JSD rose from 0.0084 in 1990 to 0.0283 in 2023 (+238%; trend p = 0.030), peaking in 2021 (+283%) with a partial 2023 pullback. Bray–Curtis dissimilarity rose +120% and the balanced panel +423%. Divergence was entirely balanced variation (share reallocation), with spectrum richness rising from 18.5 to 21.1 of 26 pathogens. Dispersion rose fastest for influenza (coefficient of variation 0.03 to 0.55) and respiratory syncytial virus (0.08 to 0.48). Within-region distance rose in every computable GBD super-region (five of seven): divergence occurs within regions, not between blocs. A country-level bootstrap confirmed the increase (+238%, 95% CI +165 to +331), and segmented trend analysis found no supported breakpoint, with divergence proceeding steadily at +4.1% per year (95% CI 2.0–6.3) rather than in stages.

**Conclusions:** National spectra are re-sorting along country-specific axes as vaccine-preventable dominance recedes at different speeds. Diversification is universal, but convergence is absent: the transition at the etiologic-spectrum level is asynchronous and path-dependent, with implications for empirical treatment policy and pathogen surveillance.

**Summary box:** *What is already known on this topic:*

- Epidemiological transition theory and the global health convergence agenda imply that national cause-of-death structures become more alike as mortality declines, but this expectation has been tested mainly at the level of mortality rates and broad cause groups, not etiologic spectra.
- Within-country (alpha) diversity of the childhood respiratory pathogen spectrum rose in 203 of 204 countries between 1990 and 2023.

*What this study adds:*

- Between-country (beta) compositional distance roughly tripled over the same period (mean pairwise Jensen–Shannon divergence +238%, 1990 to 2023), robust to distance metric, pathogen panel, COVID-19 exclusion, low-count threshold, and a fixed balanced panel.
- Divergence is driven entirely by reallocation of pathogen shares (balanced variation), is fastest for viral pathogens, and occurs within every GBD super-region, indicating that countries diversify along country-specific paths rather than toward a common spectrum.
- A non-parametric bootstrap puts a 95% confidence interval of +165% to +331% on the tripling of between-country distance between 1990 and 2023, and BIC-selected joinpoint models identify no breakpoint — divergence is steady (+4.1% per year), not staged.

## 1. Introduction

Epidemiological transition theory describes how, as mortality declines, the structure of causes of death shifts from infectious dominance toward noncommunicable disease.^1^ Although the theory was framed as a description of idealised stages rather than a law of motion, its canonical reading carries an implicit spatial expectation: countries traverse broadly similar stages, so their cause-of-death structures should become more alike as the transition advances.^2,3^ The convergence agenda in global health makes this expectation explicit at the level of mortality rates, projecting a world "converging within a generation".^4^ The empirical record at the level of life expectancy and all-cause mortality is more complicated: cross-country mortality converged through the mid-twentieth century but began to diverge again from the late 1980s, with reversal episodes concentrated in Eastern Europe and sub-Saharan Africa,^5–7^ although convergence-club analyses of mortality indicators still find country clusters converging along broadly continental lines.^8^ What has rarely been examined is the compositional layer below broad cause groups: whether the specific etiologic spectra within a single syndrome converge as countries move through the transition. The stakes for the theory are real either way. If national spectra converge as they diversify, the stage model extends naturally downward: countries follow a shared track not only in how much mortality declines but in the fine structure of what remains. If they diverge, the transition at the etiologic level is better described as asynchronous and path-dependent, with each country’s endpoint contingent on its own intervention history, and the stage imagery needs amendment. This gap is partly one of data. Compositional convergence questions require comparable, pathogen-resolved national estimates over long periods, which have only recently become available at global scale through the etiologic attribution machinery of the GBD study.^9^ Childhood respiratory infection is a natural test case. It was the dominant infectious killer of children at the start of the transition, and its mortality has fallen sharply: deaths at ages 0–19 years from the combined respiratory spectrum declined by 62.5% between 1990 and 2023 in the Global Burden of Disease Study (GBD) 2023.^9^ Over the same period, conjugate vaccines against *Streptococcus pneumoniae* and *Haemophilus influenzae* type b were introduced at different times and speeds across countries, eroding the dominance of the two historically leading pathogens unevenly.^10^ Within-country diversity of the childhood respiratory pathogen spectrum has risen nearly everywhere over the same period: in the present 204-country panel, national Shannon diversity increased in 203 of 204 countries between 1990 and 2023, and the cross-country median rose from 2.11 to 2.62. Diversification, in other words, is close to universal at the national level. Universal diversification, however, does not imply convergence. In community ecology the distinction is formal: alpha diversity measures the variety within a local community, whereas beta diversity measures compositional difference between communities.^11^ Every local community can gain diversity while the compositional distances among them widen, a pattern ecologists term differentiation, the opposite of biotic homogenisation. The same distinction applies to national pathogen spectra. Operationally, convergence would appear as shrinking pairwise compositional distance between countries over time: national spectra approaching a common template, much as mortality rates have approached common low levels. Divergence would appear as widening distance, with each country’s spectrum becoming more idiosyncratic even as all become more diverse. The two outcomes answer different questions, and the alpha-level result settles neither; indeed, in the present panel diversification was fastest where baseline diversity was lowest (convergence in slope), yet the cross-country spread of diversity widened (divergence in level), an early hint that the national trajectories are not funnelled toward a single endpoint. The question carries practical weight. Empirical treatment guidelines for childhood pneumonia are premised on a sufficiently shared pathogen spectrum that a single antibiotic algorithm is reasonable across settings;^12^ vaccine pipeline priorities and regional surveillance design make analogous assumptions about which pathogens matter where. If national spectra are in fact drifting apart, these shared templates describe a shrinking fraction of real settings.

We therefore posed three questions. First, has the between-country compositional distance of national childhood respiratory pathogen spectra increased or decreased between 1990 and 2023? Second, if distance has changed, is the change driven by reallocation of shares among a common pathogen set or by subsets of pathogens appearing and disappearing across countries? Third, which pathogens, and which geographic scales, contribute most? We addressed these questions using a national panel of pathogen-attributed respiratory deaths from GBD 2023, five timepoints spanning the vaccine era and the pandemic window, pairwise information-theoretic and abundance-based distances, and a beta-diversity decomposition transferred from community ecology. Robustness was pre-specified in both directions — convergence and divergence were each interpretable outcomes — and the analysis plan fixed the distance metrics, decomposition, low-count filter, balanced-panel check and regional stratification before results were examined. To our knowledge this is the first application of formal beta-diversity analysis to national cause-of-death spectra, and the first empirical test of compositional convergence at the etiologic level of the childhood respiratory transition.

## 2. Methods

### 2.1 Data source and spectrum definition

This is a secondary analysis of GBD 2023 (release v8352), which produces cause-specific mortality estimates for 204 countries and territories using standardised Bayesian models.^9^ Etiologically attributed lower respiratory infection (LRI) deaths were extracted at the risk–effect (rei) level for the 26 single-pathogen and residual causes of GBD 2023, for both sexes combined; attribution follows a counterfactual population-attributable-fraction (PAF) model that partitions LRI deaths across etiologies using etiologic fraction data, and the rei-level interface returns deaths (central estimates, in numbers) rather than rates or uncertainty draws.^13,14^ We aggregated the four paediatric age groups (<5, 5–9, 10–14, 15–19 years) into a single 0–19 year total and retained five timepoints: 1990, 2010, 2019, 2021 and 2023. These timepoints span the pre-vaccine baseline, the vaccine scale-up era, the immediate pre-pandemic year, the pandemic shock and the most recent post-shock year. We use 2021 rather than 2020 as the pandemic-shock timepoint: 2020 was the year of maximal non-pharmaceutical-intervention suppression of respiratory transmission, whereas 2021 is when the shock to pathogen spectra became manifest.

Three complementary causes complete the childhood respiratory infectious spectrum: tuberculosis and pertussis, which GBD models directly as causes of death rather than through PAF attribution, and COVID-19 (2021 and 2023).^13^ Because GBD causes of death are mutually exclusive and collectively exhaustive, concatenating the modules introduces no double counting. However, country-level pertussis estimates were unavailable for 1990 and 2023, and COVID-19 did not exist before 2020, so the main analysis uses the 26-pathogen LRI panel at all five timepoints. A 29-pathogen panel (adding tuberculosis, pertussis and COVID-19, with COVID-19 set to zero before 2020, which is factual) is reported as a sensitivity for 2010, 2019 and 2021.

### 2.2 Spectrum vectors and low-count filter

For each country and timepoint we formed a closed compositional vector of pathogen shares by dividing attributed deaths by the country-timepoint total. Shares derived from very small death counts are unstable: below roughly 100 deaths, single-digit changes in one pathogen’s count can swing shares by percentage points and generate spurious pairwise distances. We therefore excluded country-timepoints with fewer than 100 total attributed deaths (primary threshold). Exclusions were 67, 89, 93, 97 and 96 of 204 countries in 1990, 2010, 2019, 2021 and 2023, respectively; the rise reflects mortality decline pushing more countries, predominantly high-income and small states, below the filter. Thresholds of 50 and 200 deaths were examined in sensitivity analysis. Because a filter tied to mortality level lets the analysed sample change over time, we additionally defined a balanced panel of the 107 countries passing the 100-death filter at all five timepoints; comparing the balanced and unbalanced trends separates genuine compositional change from turnover in which countries enter the sample.

### 2.3 Between-country distance metrics

The primary metric was the Jensen–Shannon divergence (JSD) between national share vectors, computed with base-2 logarithms and bounded on [0, 1]; it is symmetric, finite everywhere (zero shares contribute zero to the entropy terms, so no pseudocounts are required), and interpretable as the information lost when two spectra are pooled.^15^ For each timepoint we computed all pairwise JSD values among included countries and summarised their distribution by the mean, median and interquartile range; with n countries this yields n(n − 1)/2 dependent pairwise values, so inference is placed on the time trend of the summary statistics rather than on individual pairs. As a sensitivity metric we used the Bray–Curtis dissimilarity, which for share vectors equals half the L1 distance and is also bounded on [0, 1].^16^ The two metrics weight dominant and rare components differently, so agreement between them indicates that the result is not metric-specific.

### 2.4 Beta-diversity decomposition

To characterise the type of compositional change, we applied the Baselga partitions. For abundance data, Bray–Curtis dissimilarity decomposes into balanced variation in abundance (countries re-allocating share among the same components) and abundance gradients (one country’s components forming a strict subset of another’s).^17^ For closed share vectors, both share totals equal one by construction, so the gradient component is identically zero; we report this as a mathematical property and therefore pair the abundance partition with an incidence-based partition. Converting shares to presence (share ≥0.5%) and absence, we decomposed Jaccard dissimilarity into turnover and nestedness components,^18^ and tracked mean spectrum richness (number of pathogens present at ≥0.5% share) per timepoint. The 0.5% presence threshold is arbitrary but conservative: it is high enough that a pathogen must carry a non-trivial mortality share to count as present, and low enough that the long tail of the spectrum remains visible.

### 2.5 Per-pathogen dispersion and regional stratification

For each pathogen and timepoint we computed the cross-country coefficient of variation (CV, standard deviation divided by mean) and interquartile range of its share; the CV is scale-free, so pathogens with small mean shares can be compared with dominant ones. Countries were mapped to the seven GBD super-regions using the standard GBD location hierarchy, and mean pairwise JSD was computed within each super-region (where at least five countries passed the filter) and between country pairs spanning different super-regions, to test whether divergence reflects within-region differentiation or regional blocs drifting apart.

### 2.6 Statistics and reporting

Temporal trend was assessed by ordinary least squares regression of mean pairwise distance on calendar year across the five timepoints, reporting the slope, Pearson correlation and two-sided p value; with five points the test has three residual degrees of freedom and is interpreted as descriptive evidence of direction rather than confirmatory inference. Because pairwise distances are not independent, the primary regression was accompanied by an exact permutation test over the 120 orderings of the five timepoint labels. Several such trend tests were performed on the same underlying distance summaries (the primary JSD, Bray–Curtis and balanced-panel series); these tests are correlated, no adjustment for multiple comparisons was made, and the marginal p = 0.030 of the primary series should be read with corresponding caution, the strength of evidence resting on effect size and consistency across specifications rather than on any single p value. All other analyses are descriptive. Analyses used Python (pandas, NumPy, SciPy) with two-sided alpha 0.05. The study uses publicly available, de-identified, aggregate modelled estimates and was exempt from institutional ethics review.

Three analyses were added in this revision. First, to test whether divergence accelerated at identifiable times, segmented log-linear (joinpoint) models were fitted to the five-timepoint series of mean pairwise JSD (primary and balanced-panel) and mean Bray–Curtis dissimilarity, with the number and location of joinpoints selected by the Bayesian information criterion; because every segment was required to contain at least three observations (adjacent joinpoints at least two years apart), five timepoints admit at most one breakpoint, at 2019. Second, to quantify the sensitivity of the incidence-based partition to the arbitrary 0.5% presence threshold, the Jaccard turnover–nestedness decomposition and mean richness were recomputed at thresholds of 0.25%, 1% and 2% of spectrum share. Third, a non-parametric bootstrap (2,000 resamples of countries with replacement within each timepoint; seed 42) gave 95% confidence intervals for the 1990 to 2023 change in mean pairwise JSD in absolute and percentage terms. Full segment tables, the threshold-sensitivity grid and the bootstrap summary accompany this revision as CSV files with the reproducible script (Supplementary_Joinpoint_DivergingSpectra.csv, sensitivity_presence_threshold_DivergingSpectra.csv, bootstrap_jsd_change_DivergingSpectra.csv; code/joinpoint_robustness_divergingspectra.py).

## 3. Results

### 3.1 Between-country compositional distance tripled between 1990 and 2023

In 1990 national spectra were near-identical: the mean pairwise JSD among the 137 included countries was 0.0084 on a [0, 1] scale (median 0.0051), reflecting the shared dominance of *S. pneumoniae* (48.6% mean share) and broad residual categories. Part of this early homogeneity is likely to reflect covariate-driven smoothing of the GBD attribution models in the data-sparsest year, so the magnitude of the subsequent increase is interpreted conservatively (see Limitations). Distance then rose at every successive timepoint to a 2021 peak of 0.0320 (+283% relative to 1990), before a partial pullback to 0.0283 in 2023 (Table 1, Figure 1). The 2023 level remains more than threefold the 1990 level (+238%; linear trend over the five timepoints: slope 0.00068 per year, r = 0.91, p = 0.030; exact permutation p = 0.025). The median pairwise distance tracked the mean (0.0051 to 0.0240), indicating a shift of the whole distribution rather than a tail phenomenon. The Bray–Curtis sensitivity metric showed the same pattern: mean dissimilarity rose from 0.066 to 0.145 (+120%; r = 0.95, p = 0.014). Absolute JSD values are small on their [0, 1] scale; we therefore report relative change alongside absolute levels throughout.

**Figure 1.**
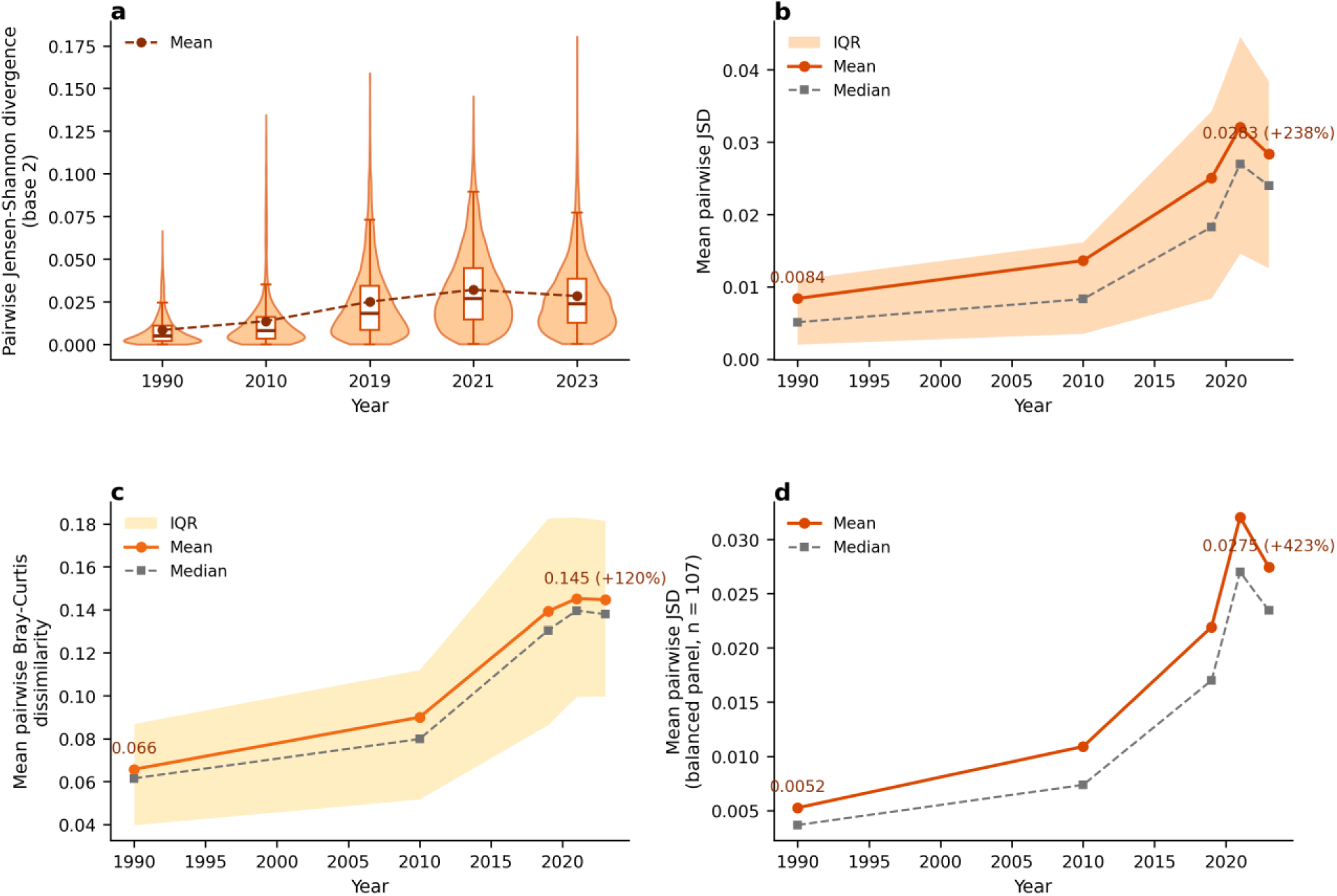
Pairwise between-country Jensen–Shannon divergence of national childhood respiratory pathogen spectra, 1990–2023. *(a) Distribution of all pairwise JSD values among included countries at each of five timepoints (violins with box plots and mean trace; 26-pathogen panel, 100-death filter). (b) Mean pairwise JSD with interquartile band, 0.0084 (1990) to 0.0283 (2023; +238%); the 2021 pandemic-window peak (0.0320) and partial 2023 pullback are visible. (c) Bray–Curtis sensitivity metric, mean with interquartile band (0.066 to 0.145; +120%). (d) Balanced panel of the 107 countries passing the filter at all five timepoints (0.0052 to 0.0275; +423%)*.

**Table 1.** Pairwise between-country compositional distance of national childhood respiratory pathogen spectra (26 pathogens, ages 0–19 years), 1990–2023.

| **Year** | **Countries, n** | **JSD mean** | **JSD median** | **JSD IQR** | **Bray–Curtis mean** | **Bray–Curtis median** |
|---|---|---|---|---|---|---|
| 1990 | 137 | 0.0084 | 0.0051 | 0.0020–0.0110 | 0.066 | 0.061 |
| 2010 | 115 | 0.0136 | 0.0083 | 0.0035–0.0162 | 0.090 | 0.080 |
| 2019 | 111 | 0.0250 | 0.0183 | 0.0084–0.0343 | 0.139 | 0.130 |
| 2021 | 107 | 0.0320 | 0.0270 | 0.0146–0.0446 | 0.145 | 0.140 |
| 2023 | 108 | 0.0283 | 0.0240 | 0.0126–0.0384 | 0.145 | 0.138 |

JSD, Jensen–Shannon divergence (base 2, bounded [0, 1]); IQR, interquartile range. Change 1990 to 2023: JSD mean +238% (trend p = 0.030); Bray–Curtis mean +120% (p = 0.014).

Two additions firm up the trend inference. Segmented (joinpoint) models selected by the Bayesian information criterion retained zero joinpoints for the primary JSD series, the balanced-panel series and the Bray–Curtis series alike: under the constraint of at least three observations per segment, five timepoints admit at most one breakpoint (2019), and the 2021 peak with its 2023 pullback did not justify one. The single admissible breakpoint at 2019 was evaluated explicitly and was rejected for all three series (ΔBIC = +2.8, +2.6 and +3.1 for the primary, balanced-panel and Bray–Curtis series, respectively; positive ΔBIC favours the no-joinpoint model). Divergence is therefore best described as a single steady log-linear rise—+4.1% per year (95% CI 2.0–6.3) for mean pairwise JSD, +5.5% (3.0–8.1) on the balanced panel and +2.6% (1.4–3.8) for Bray–Curtis—rather than as staged or purely pandemic-driven acceleration. A non-parametric bootstrap over countries (2,000 resamples within timepoints) put a 95% confidence interval of +165% to +331% on the +238% rise in mean pairwise JSD between 1990 and 2023 (absolute change +0.0200, 95% CI 0.0155–0.0244), complementing the exact permutation test by quantifying sampling uncertainty in the magnitude of change.

### 3.2 Divergence is robust to metric, panel composition, pathogen set, COVID-19 and count threshold

The result did not depend on which countries entered the sample. On the balanced panel of 107 countries passing the low-count filter at all five timepoints, mean pairwise JSD rose from 0.0052 to 0.0275 (+423%; p = 0.038), steeper than in the unbalanced sample, so divergence is not an artefact of changing sample composition (Table 2). Extending the panel to 29 pathogens (adding tuberculosis, pertussis and COVID-19) for 2010–2021 preserved the direction with a steeper slope: mean JSD rose from 0.056 (2010) through 0.067 (2019) to 0.103 (2021), at a higher absolute level because tuberculosis and pertussis add cross-country heterogeneity. COVID-19 amplified but did not drive the pandemic-era peak: excluding it, the 2021 mean JSD on the 28-pathogen panel was 0.072 versus 0.103 with it (+43% amplification), and the 2010 to 2021 trend was identical in direction (0.056 to 0.067 to 0.072). Low-count thresholds of 50, 100 and 200 deaths all yielded increases of +238% to +255% between 1990 and 2023 (Figure 5).

**Table 2.**
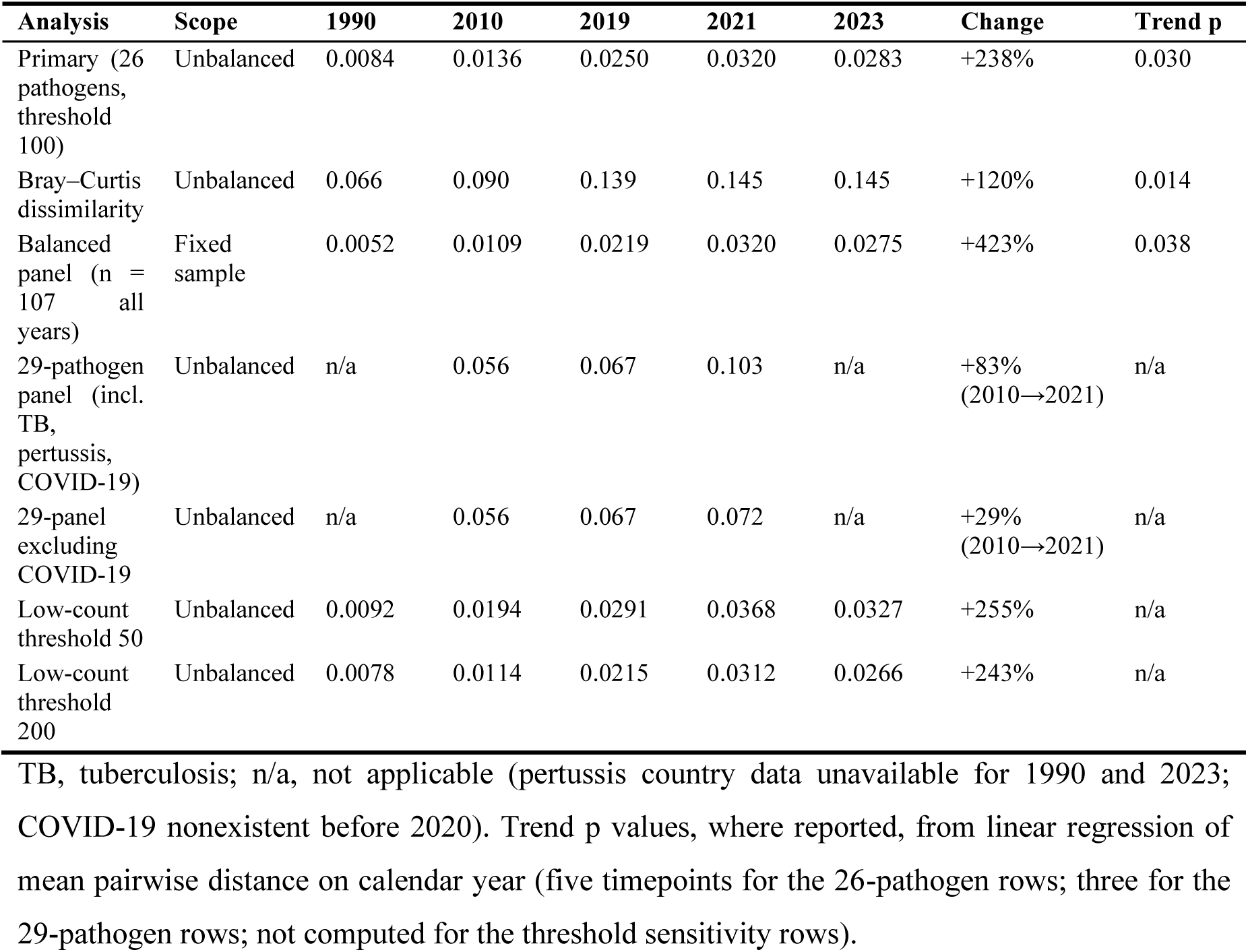
Robustness of the divergence trend (mean pairwise JSD)

| Analysis | Scope | 1990 | 2010 | 2019 | 2021 | 2023 | Change | Trend p |
|---|---|---|---|---|---|---|---|---|
| Primary (26 pathogens, threshold 100) | Unbalanced | 0.0084 | 0.0136 | 0.0250 | 0.0320 | 0.0283 | +238% | 0.030 |
| Bray–Curtis dissimilarity | Unbalanced | 0.066 | 0.090 | 0.139 | 0.145 | 0.145 | +120% | 0.014 |
| Balanced panel (n = 107 all years) | Fixed sample | 0.0052 | 0.0109 | 0.0219 | 0.0320 | 0.0275 | +423% | 0.038 |
| 29-pathogen panel (incl. TB, pertussis, COVID-19) | Unbalanced | n/a | 0.056 | 0.067 | 0.103 | n/a | +83%<br>(2010→2021) | n/a |
| 29-panel excluding COVID-19 | Unbalanced | n/a | 0.056 | 0.067 | 0.072 | n/a | +29%<br>(2010→2021) | n/a |
| Low-count threshold 50 | Unbalanced | 0.0092 | 0.0194 | 0.0291 | 0.0368 | 0.0327 | +255% | n/a |
| Low-count threshold 200 | Unbalanced | 0.0078 | 0.0114 | 0.0215 | 0.0312 | 0.0266 | +243% | n/a |

TB, tuberculosis; n/a, not applicable (pertussis country data unavailable for 1990 and 2023; COVID-19 nonexistent before 2020). Trend p values, where reported, from linear regression of mean pairwise distance on calendar year (five timepoints for the 26-pathogen rows; three for the 29-pathogen rows; not computed for the threshold sensitivity rows).

### 3.3 Divergence is share reallocation, with a nestedness signature in incidence space

The Baselga abundance partition attributed 100% of quantitative divergence at every timepoint to balanced variation in abundance, the turnover analogue: countries are re-allocating shares among a common pathogen set (Table 3, Figure 2). The abundance-gradient component was identically zero, a mathematical property of closed share vectors rather than an empirical finding, so we verified the structural pattern in incidence space. There, total Jaccard dissimilarity was roughly stable (0.052 in 1990; 0.059 in 2023; a 2021 excursion to 0.089) while mean spectrum richness rose from 18.5 to 21.1 of 26 pathogens. Because richer spectra tend to contain poorer ones, nestedness accounted for the majority of incidence dissimilarity (68% in 2023 versus 57% in 1990; turnover fraction 32% versus 43%). The turnover fraction was non-monotonic across the intermediate timepoints, however (11% in 2010 against 32–43% in the other years), so the nestedness majority is an endpoint pattern rather than a stable ordering and is interpreted descriptively. The combined picture is consistent: countries are gaining components almost universally, yet the proportional composition of the enriched spectra is increasingly country-specific. Re-running the partition at presence thresholds of 0.25%, 1% and 2% (base 0.5%) left the qualitative pattern intact: mean richness rose between 1990 and 2023 at every threshold (21.2 to 23.1, 15.9 to 17.7 and 11.2 to 13.2 of 26 pathogens, versus 18.5 to 21.1 at base), and nestedness remained the majority component of 2023 incidence dissimilarity at thresholds up to 1% (turnover fraction 2.5%, 31.8% and 49.3% at 0.25%, 0.5% and 1%), tipping to a narrow turnover majority (54.1%) only at the strict 2% cut-off. The nestedness signature is therefore robust across the plausible threshold range, even though the exact turnover–nestedness split is threshold-dependent (sensitivity_presence_threshold_DivergingSpectra.csv).

**Figure 2.**
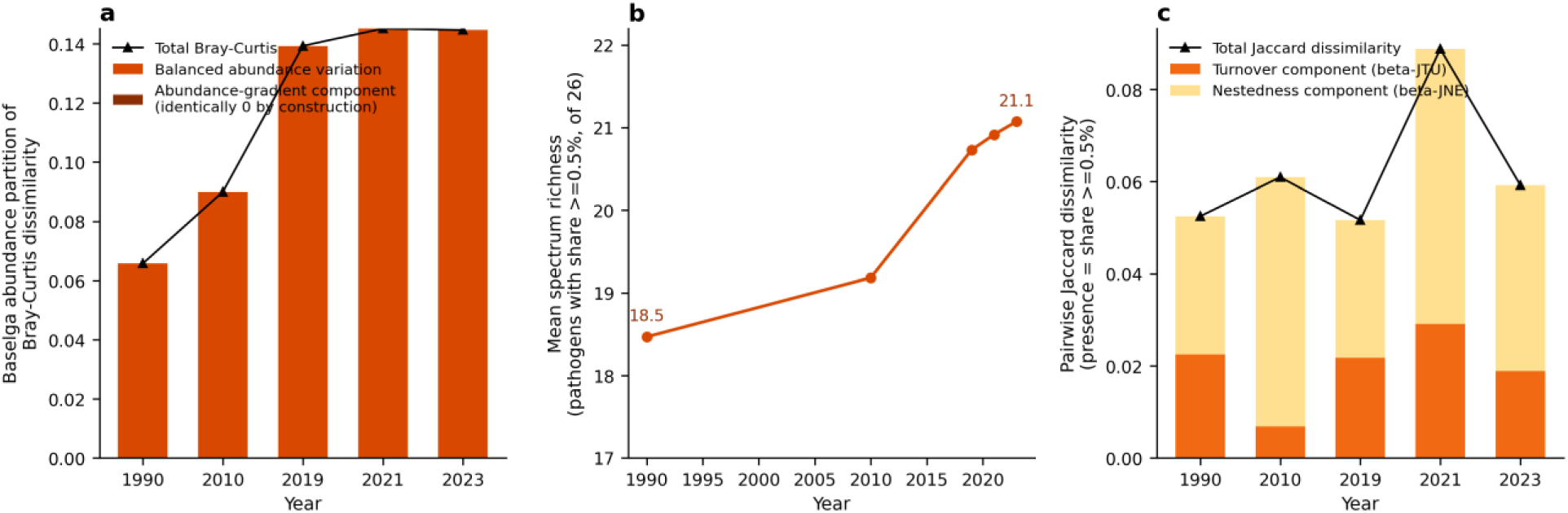
Beta-diversity decomposition of between-country dissimilarity, 1990–2023. *(a) Baselga abundance partition of Bray–Curtis dissimilarity into balanced variation and abundance-gradient components; the 100%/0% split is an identity by construction for closed share vectors, shown for completeness rather than as an empirical result. (b) Mean spectrum richness (pathogens with share ≥0.5%, of 26), rising from 18.5 to 21.1. (c) Jaccard partition into turnover and nestedness components with the total Jaccard dissimilarity series*.

**Table 3.**
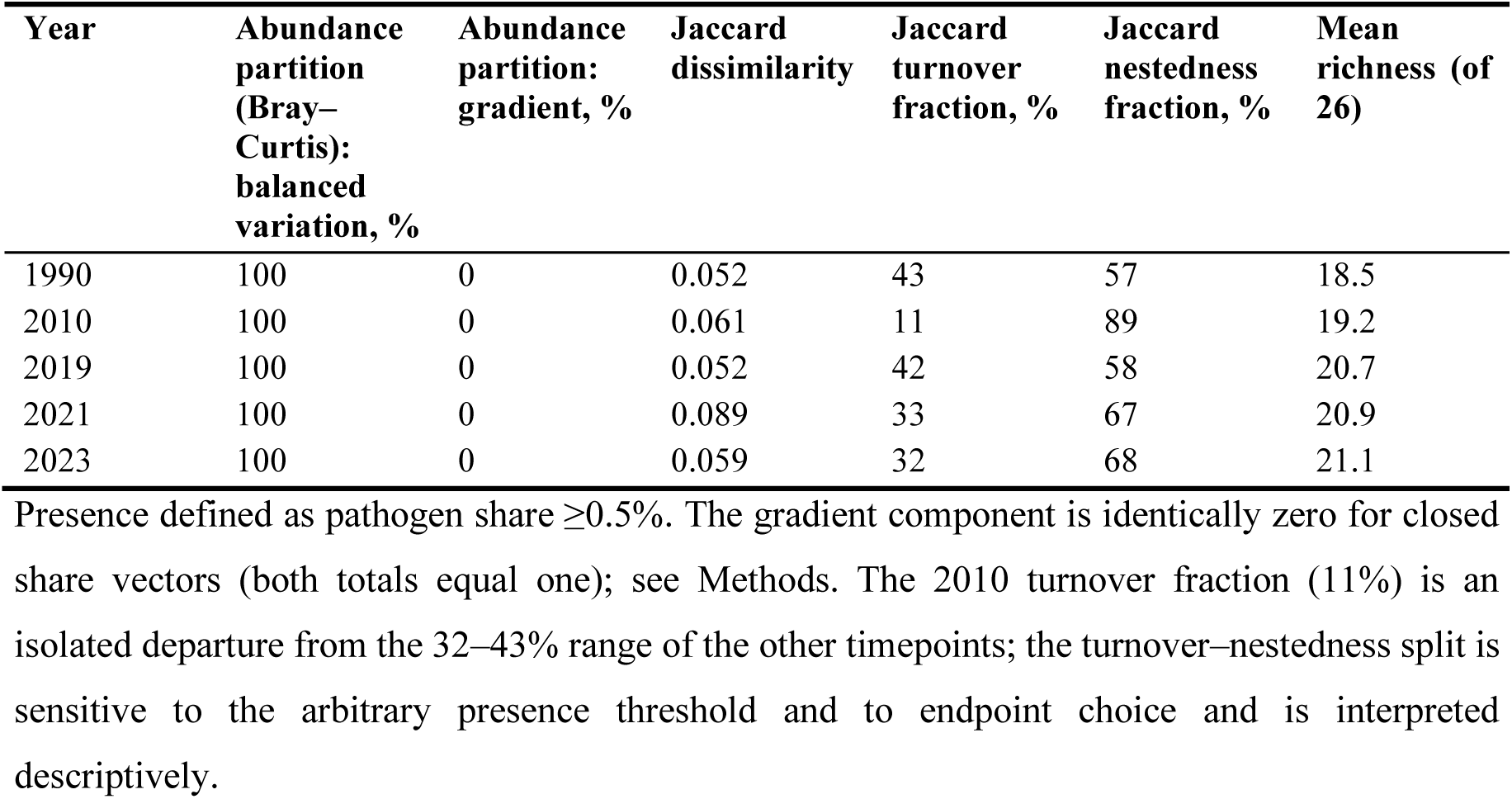
Beta-diversity decomposition of between-country dissimilarity, 1990–2023.

### 3.4 Viral pathogens show the fastest cross-country dispersion

Fifteen of 26 pathogens showed rising cross-country CV of share between 1990 and 2023 (Table 4, Figure 3). Dispersion rose fastest for the two major respiratory viruses: influenza from 0.03 to 0.55 and respiratory syncytial virus (RSV) from 0.08 to 0.48, despite broadly stable mean shares, meaning that influenza and RSV mortality shares were nearly uniform across countries in 1990 a uniformity probably exaggerated by covariate-driven model smoothing in a data-sparse year but differ widely in 2023. *S. pneumoniae* itself dispersed markedly (0.06 to 0.26) as its mean share fell from 48.6% to 29.5%, consistent with uneven pacing of the post-pneumococcal transition. *Mycoplasma*, other gram-negative bacteria, *Pseudomonas aeruginosa* and *Escherichia coli* dispersed from low bases. Several high-CV opportunistic or rare causes were stable or declining (*Klebsiella pneumoniae* 0.19 to 0.18; *Staphylococcus aureus* 0.32 to 0.31; *Aspergillus* spp. 0.39 to 0.33). *Legionella* spp. was the counterpoint: the highest CV in the panel at every timepoint, yet falling from 2.86 to 2.04 — the largest absolute dispersion narrowed rather than widened.

**Figure 3.**
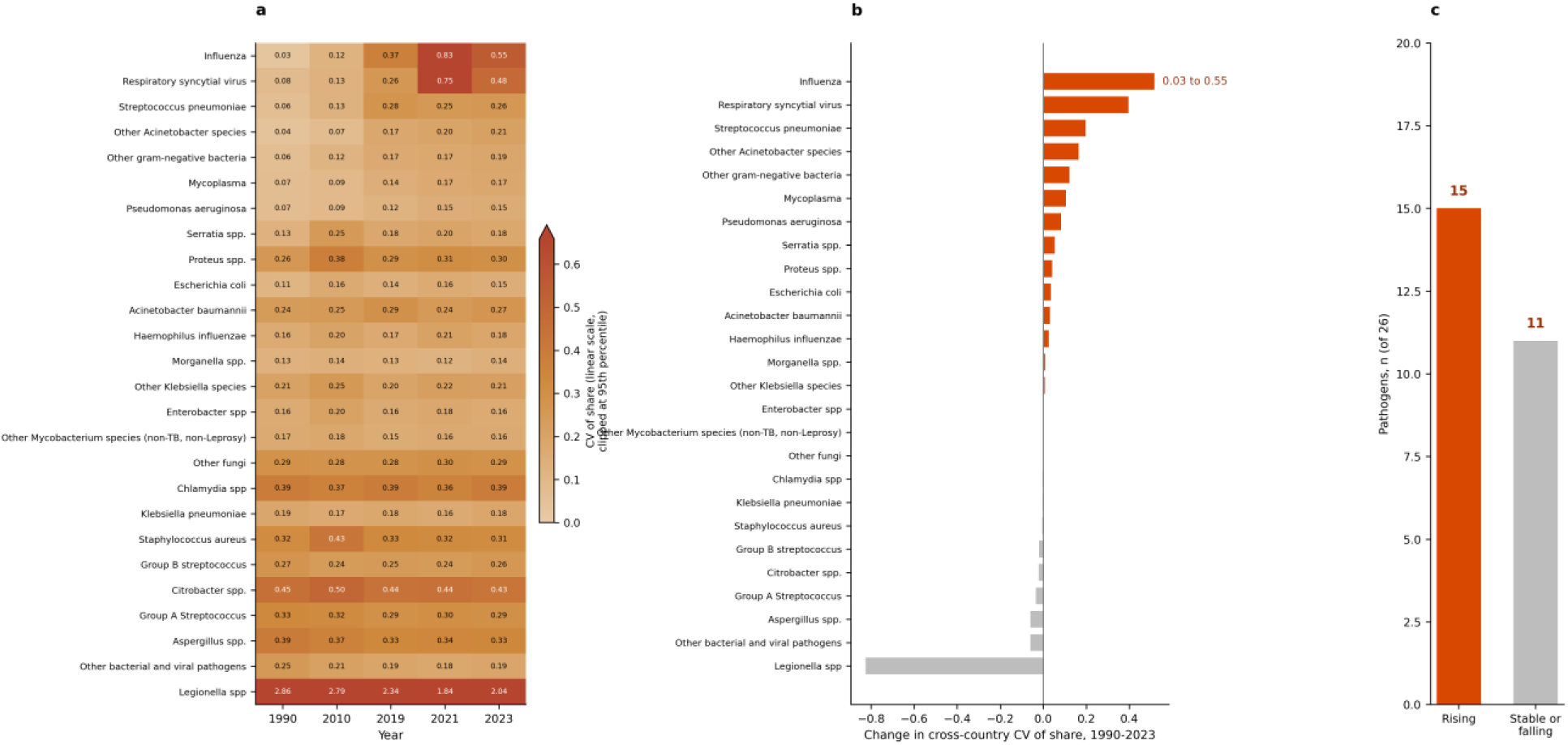
Cross-country dispersion of pathogen shares. *(a) Heat map of the coefficient of variation (CV) of national share for each of 26 pathogens across the five timepoints, ordered by the 1990–2023 change in CV (largest increase first); cell annotations are raw CV values and colour is on a linear scale from 0, clipped at the 95th percentile of all cells (0.66), so the maximum colour denotes CV ≥0.66, including Legionella spp. (CV 1.84–2.86). (b) Ranked 1990–2023 change in CV per pathogen; dispersion rose fastest for influenza (0.03 to 0.55) and RSV (0.08 to 0.48). (c) Count of pathogens with rising (15) versus stable or falling (11) cross-country CV*.

**Table 4.**
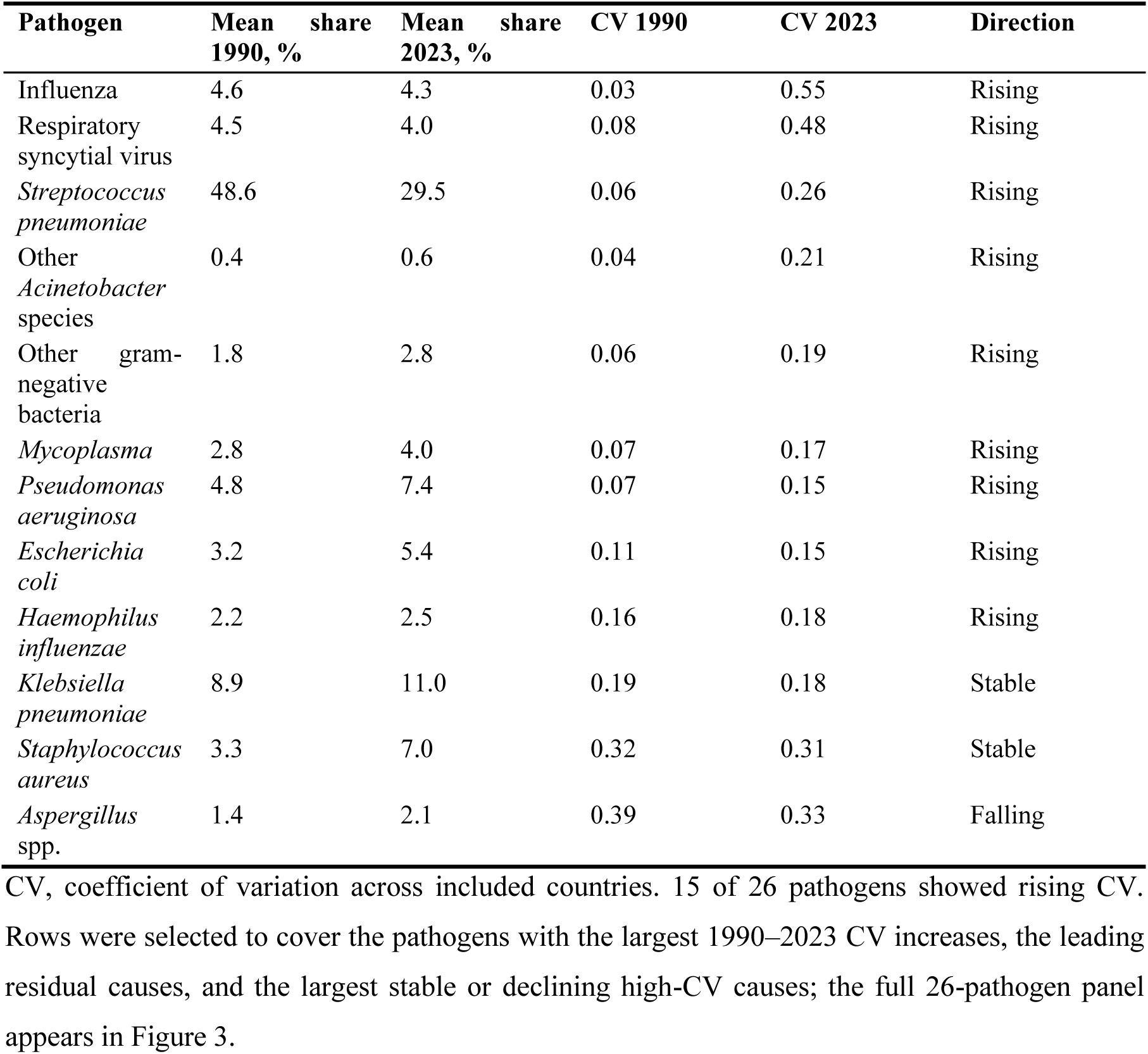
Cross-country dispersion (coefficient of variation) of pathogen shares, selected pathogens, 1990 versus 2023.

### 3.5 Divergence occurs within every super-region

Mean within-region pairwise JSD rose between 1990 and 2023 in all five super-regions with computable samples (Table 5, Figure 4): 3.5-fold in Southeast Asia, East Asia and Oceania; 4.4-fold in Central Europe, Eastern Europe and Central Asia; 5.6-fold in Latin America and the Caribbean; 6.0-fold in North Africa and the Middle East; and 9.0-fold in Sub-Saharan Africa. Mean JSD between country pairs spanning different super-regions rose 3.2-fold (0.0096 to 0.0310), nearly matching the global 3.4-fold rise. Divergence is therefore a within-region phenomenon, not a story of regional blocs drifting apart. The two remaining super-regions also rose over their shorter computable windows (South Asia 0.0009 to 0.0020 and the high-income super-region 0.0064 to 0.0337 between 1990 and 2010), so within-region divergence held in all seven super-regions wherever it could be measured. South Asia and the high-income super-region could not be assessed in later years (four and three included countries in 2023, respectively), itself a consequence of mortality decline in those settings.

**Figure 4.**
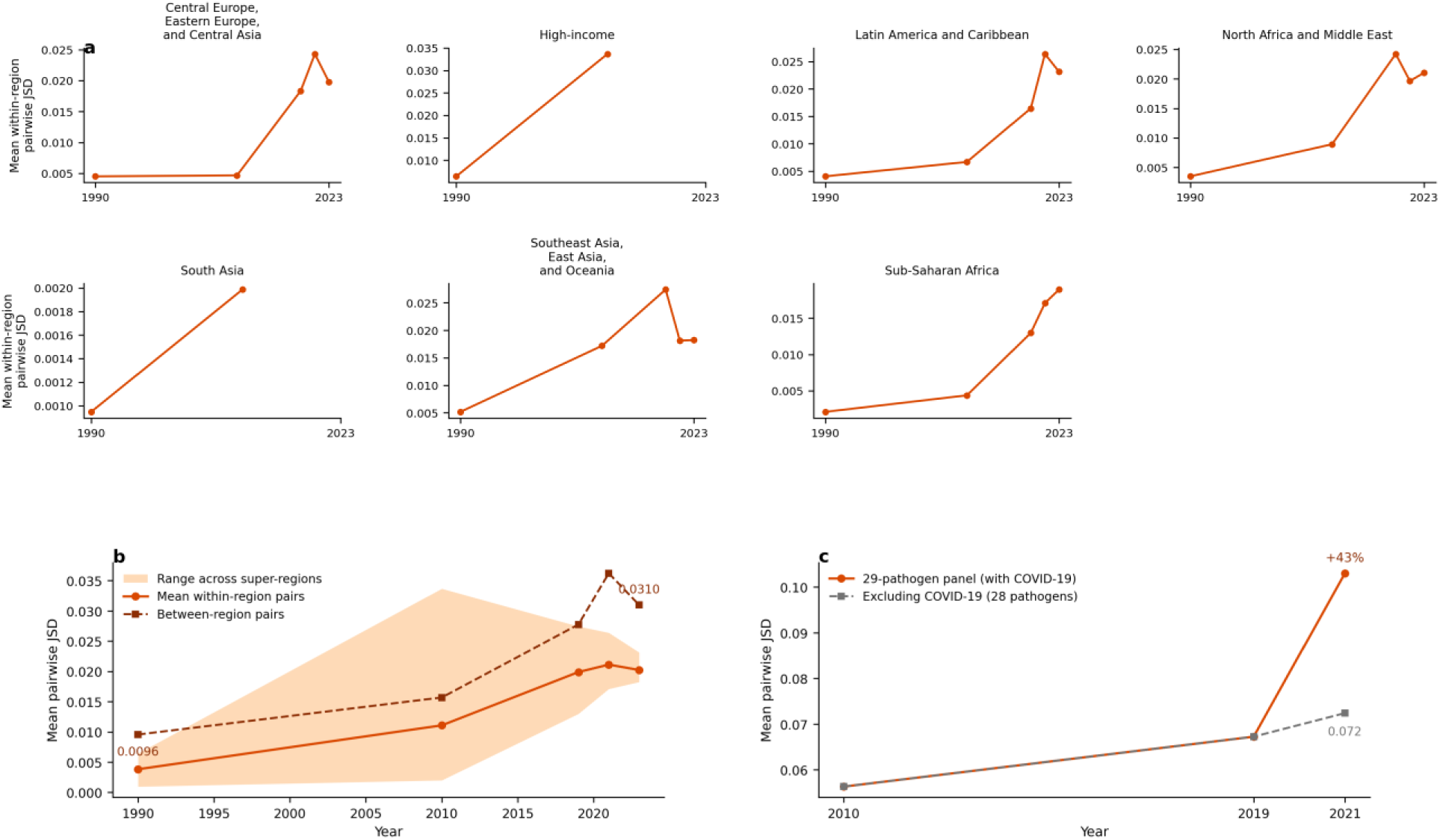
Within-region divergence across GBD super-regions. *(a) Mean within-region pairwise JSD at each timepoint, shown as small multiples for all seven super-regions (South Asia and the high-income super-region are computable only through 2010)*. *(b) Mean within-region pairs and the range across super-regions versus between-region pairs (0.0096 to 0.0310): divergence is a within-region phenomenon. (c) COVID-19 sensitivity on the extended panel, 2010–2021: mean pairwise JSD with the 29-pathogen panel (0.056 to 0.067 to 0.103) versus the 28-pathogen panel excluding COVID-19 (0.056 to 0.067 to 0.072); COVID-19 amplifies the 2021 peak by +43% but does not drive the trend*.

**Figure 5.**
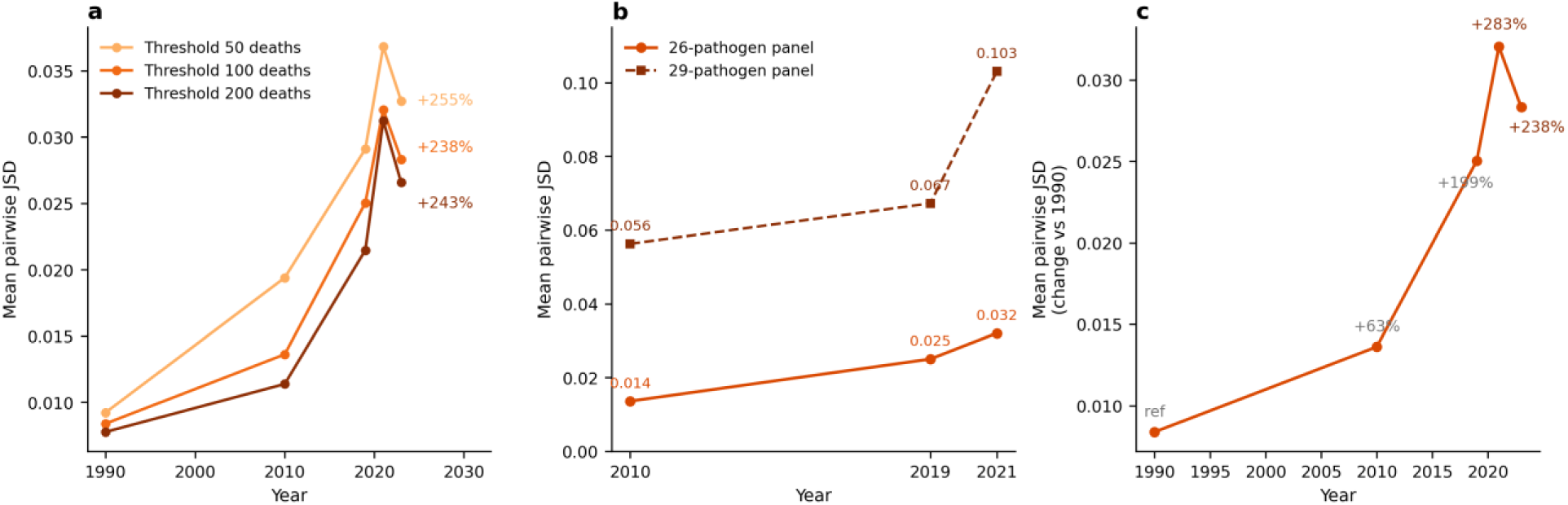
Robustness overview. *(a) Mean pairwise JSD under low-count thresholds of 50, 100 and 200 deaths; the 1990–2023 increase is +255%, +238% and +243%, respectively. (b) Twenty-six-versus 29-pathogen panels (adding tuberculosis, pertussis and COVID-19), 2010–2021. (c) Primary series annotated as percentage change relative to 1990: +63% (2010), +199% (2019), +283% (2021 peak) and +238% (2023)*.

**Table 5.** Mean pairwise JSD within GBD super-regions and between super-region pairs, 1990–2023.

| **Super-region** | **Countries 1990, n** | **Countries 2023, n** | **JSD 1990** | **JSD 2010** | **JSD 2019** | **JSD 2021** | **JSD 2023** | **Fold change** |
|---|---|---|---|---|---|---|---|---|
| Sub-Saharan Africa | 44 | 44 | 0.0021 | 0.0044 | 0.0130 | 0.0171 | 0.0190 | ×9.0 |
| Latin America and Caribbean | 19 | 16 | 0.0041 | 0.0067 | 0.0165 | 0.0264 | 0.0231 | ×5.6 |
| North Africa and Middle East | 18 | 16 | 0.0035 | 0.0089 | 0.0242 | 0.0196 | 0.0210 | ×6.0 |
| Central Europe, Eastern Europe, Central Asia | 23 | 12 | 0.0045 | 0.0047 | 0.0183 | 0.0243 | 0.0197 | ×4.4 |
| Southeast Asia, East Asia and Oceania | 16 | 13 | 0.0052 | 0.0172 | 0.0274 | 0.0181 | 0.0182 | ×3.5 |
| South Asia | 5 | 4 | 0.0009 | 0.0020 | n/c | n/c | n/c | n/c |
| High-income | 12 | 3 | 0.0064 | 0.0337 | n/c | n/c | n/c | n/c |
| Between-region pairs | n/a | n/a | 0.0096 | 0.0157 | 0.0277 | 0.0362 | 0.0310 | ×3.2 |

JSD, Jensen–Shannon divergence; n/c, not computed (fewer than five included countries); n/a, not applicable (between-region pairs have no region membership). Fold change 1990 to 2023.

## 4. Discussion

### 4.1 Principal findings

Between 1990 and 2023, the compositional distance among national childhood respiratory pathogen spectra roughly tripled, whether measured by Jensen–Shannon divergence (+238%) or Bray–Curtis dissimilarity (+120%), and whether or not sample composition, pathogen panel, COVID-19 and count thresholds were varied. The distance distribution shifted as a whole rather than at the extremes. Divergence peaked during the pandemic year 2021 (+283%) and partially receded by 2023, but the 2023 level remained more than threefold the 1990 baseline. Decomposition showed the divergence to be entirely balanced variation: countries re-allocating shares among a common, and slowly expanding, pathogen set. Dispersion rose fastest for the major respiratory viruses and for pneumococcus itself, and divergence occurred within every assessable super-region. Set against the near-universal rise in within-country diversity over the same period (203 of 204 countries gained Shannon diversity), the picture is consistent: national spectra are diversifying almost everywhere, and they are doing so along different paths. Because the near-uniform 1990 baseline is partly a smoothed model output (Limitations), we read the increase conservatively as at least a doubling of between-country distance.

### 4.2 Implications for epidemiological transition theory

These results add a second layer to the transition narrative. At the level of mortality rates and broad cause groups, the transition literature has documented convergence followed by divergence, driven by reversal episodes in particular regions.^5–7^ At the etiologic level studied here, we find no convergence phase at all within the 1990–2023 window: national spectra began the period near-identical, dominated everywhere by pneumococcus and residual categories, and have been differentiating ever since. The mechanism is plausibly the asynchronous, path-dependent unwinding of that shared starting point. Conjugate vaccine introduction varied by more than two decades across countries,^10^ and national third-dose pneumococcal conjugate vaccine coverage still spans from nil in countries yet to introduce it to near-universal levels elsewhere,^19^ so as vaccine-preventable dominance recedes at different speeds, the revealed spectrum re-sorts along country-specific axes: viral-dominated in some settings, gram-negative or opportunistic mixes in others, depending on local vaccine coverage, antimicrobial pressure and health-care exposure. The 2021 peak sharpens the point. A global shock that struck every country simultaneously did not homogenise national spectra; it amplified their differences by 43% on top of the secular trend, because the shock interacted with already-divergent structures (asynchronous disruption and resurgence of respiratory viruses is documented in incidence surveillance).^20^ The partial 2023 pullback suggests a reversible shock component superimposed on a persistent secular component, and the 2019 value (0.0250), measured before any pandemic effect, already sits threefold above 1990, so the trend does not depend on the pandemic window. If this reading is correct, the "post-pneumococcal diaspora" is not a transitional phase on the way to a new common spectrum; it is the transition, proceeding at different speeds along different paths. Transition theory’s stage imagery, in which countries follow one another along a shared track, fits the etiologic layer poorly: the stages were defined by the order in which broad cause groups recede, but the residual composition left behind by that recession is contingent on national intervention histories and health-system structures, and contingency compounds over time. The within-region pattern reinforces this interpretation. Divergence is not driven by regional blocs drifting apart; neighbouring countries with similar starting points are acquiring dissimilar spectra, which is what path dependence looks like at this scale.

### 4.3 Clinical and policy implications

Empirical management of childhood pneumonia assumes that a shared pathogen spectrum justifies shared antibiotic algorithms.^12^ That premise is weakening between countries. A spectrum distance that has at least doubled raises the possibility that any single empirical regimen may increasingly mismatch local spectra, in setting-specific ways. This inference is necessarily indirect: part of the divergence is driven by viral shares that antibacterial algorithms do not target, the pathogen mix of fatal infections need not match that of presenting clinical pneumonia, and empirical choices in practice hinge on local resistance patterns, which we did not measure. The per-pathogen results make the point concrete: influenza and RSV shares were nearly uniform across countries in 1990 but now vary several-fold to more than tenfold more widely, so the viral contribution to severe pneumonia presentations, and hence the likely yield of antibacterial-first management, differs increasingly by setting. Three practical consequences follow. First, regional and national etiologic surveillance gains value relative to global averages: as between-country heterogeneity grows, pooled estimates describe fewer individual countries well, and the case strengthens for sentinel etiologic testing networks feeding national guideline committees. Second, intervention portfolios need local compositional information. As single-pathogen strategies address a shrinking share of the spectrum almost everywhere, platform interventions (oxygen, antimicrobial access, referral) gain relative weight; the present result adds that the identity of the leading residual pathogens differs increasingly by country, so platforms must be stocked and calibrated locally. Third, vaccine development and introduction priorities, for example for RSV, will have heterogeneous marginal value across countries whose spectra have already diverged, which argues for country-stratified rather than globally pooled estimates of vaccine-preventable burden in investment cases.

### 4.4 A beta-diversity framework for monitoring etiologic transitions

Methodologically, the study demonstrates that the alpha/beta apparatus of community ecology^11,17,18^ transfers directly to population-level pathogen spectra and answers a question that alpha-diversity reporting alone cannot pose. The present study illustrates the two-layer reporting standard we would propose for etiologic-transition monitoring: alpha diversity (is the national spectrum diversifying?), beta diversity (are national spectra converging?), and decomposition (is change driven by share reallocation or by components appearing and disappearing?). Jensen–Shannon divergence is a convenient primary metric because it is bounded, symmetric and information-theoretically interpretable;^15^ the Baselga partitions then discipline interpretation by separating turnover-like from nestedness-like structure.^17,18^ Two features of the present application may be worth standardising. First, the balanced-panel check proved essential: in any monitoring system where units drop below a reporting threshold as the underlying burden falls, unbalanced samples can manufacture or mask trends, and the fixed-sample comparison should be routine. Second, the distinction between balanced variation and nestedness has a direct policy translation: reallocation within a shared pathogen set calls for recalibration of existing tools, whereas genuine turnover of the pathogen set would call for new tools. The same framework could monitor spectra of antimicrobial resistance, serotype replacement after vaccine introduction, or any multi-cause mortality structure in which compositional change, not just level change, is of interest. We have not modelled why trajectories differ; geographic and ecological covariates — island-state status, latitude and the distance decay of compositional similarity — are the natural next driver analysis, one that must reckon with the low-count filter, which already excludes most small island states from the panel.

### 4.5 Relation to the convergence literature

Our finding appears to contradict the convergence rhetoric in global health,^4^ but the discrepancy is one of level and metric, not of fact. Convergence claims concern mortality levels (under-five mortality rates, life expectancy gaps), where catch-up is real; the documented late-twentieth-century divergence likewise concerns levels,^5–7^ and the convergence clubs identified by clustering country mortality indicators are clubs of levels, not of compositions.^8^ Compositional structure is a different object. Two countries can have identical and rapidly falling pneumonia mortality while the mix of pathogens killing their remaining cases grows steadily apart — precisely what the balanced panel shows. Direct precedents for compositional comparison are scarce: cause-of-death structure has been compared across countries mainly through broad groupings (communicable versus noncommunicable versus injury), at which aggregation level the transition’s shared direction dominates and similarity necessarily rises. The closest precedents are within-country (alpha-level) analyses of cause-of-death diversity — in low-mortality countries^21^ and globally across 204 countries, where diversity trends themselves differed by region^22^ — analogues of the within-country diversification documented here rather than tests of between-country convergence. At the 26-pathogen resolution studied here, the shared direction of decline drops out of the measure, because shares are normalised within country and year; what remains is pure compositional difference, and it grows. Even within the alpha layer, the present panel shows convergence in slope (diversification was fastest where baseline diversity was lowest) but divergence in level (the cross-country spread of Shannon diversity widened). The layered picture — converging rates, diverging diversity levels and diverging compositions — is more coherent than any single convergence verdict, and it suggests that "convergence" should be specified by layer before it is asserted.

### 4.6 Limitations

Several limitations qualify the findings. First, GBD etiologic attributions are modelled estimates derived from counterfactual population-attributable fractions, not etiologic testing of individual deaths;^13,14,23^ pathogen shares in low-data settings borrow strength from covariates and regional patterns, which could attenuate or inflate measured divergence in ways we cannot isolate. This caveat weighs most heavily on the 1990 baseline. Eleven of the 26 pathogens were newly modelled in GBD 2023,^14^ and national estimates for the earliest, data-sparsest years lean more heavily on covariate-driven smoothing, so part of the near-uniformity of 1990 spectra may be a modelling artefact and the headline +238% may partly reflect the growth of information content over time rather than compositional change alone. Two sensitivity results partially address this concern: on the balanced panel of 107 continuously included countries the increase was steeper, not weaker (+423%), and under the stricter 200-death threshold, which retains only higher-burden settings, the increase remained +243% (+255% under the 50-death threshold). We therefore retain the directional conclusion but describe the magnitude as at least a twofold increase rather than a precisely estimated threefold one. Second, uncertainty was not propagated: the rei-level interface returns central estimates only, so pairwise distances, trend statistics and p values carry no uncertainty bands. GBD 2023 uncertainty intervals around pathogen-attributed deaths are wide in low-data settings,^14^ and because pairwise distance is a nonlinear function of shares, the reported p values overstate precision to a degree we cannot quantify. Third, the low-count filter progressively excludes countries as mortality declines (67 of 204 excluded in 1990; 96 in 2023), so the later sample is predominantly low- and middle-income; the balanced panel shows the trend is stronger, not weaker, within a fixed sample, but our estimates do not describe high-income spectra in later years and the within-region analysis could not assess two super-regions by 2023. Fourth, five timepoints do not constitute a continuous series; the trend test has three residual degrees of freedom, and p = 0.030 should be read as modest statistical evidence resting on a large and consistent relative change, corroborated across specifications. Fifth, the 2021 timepoint contains the pandemic shock; we quantified and separated the COVID-19 contribution, but pandemic-era attribution is unusually uncertain. Sixth, shares form a closed composition, which induces mathematical coupling: a fall in pneumococcal share mechanically raises other shares, and the abundance-gradient component of the decomposition is zero by construction. We mitigated this by pairing the abundance partition with an incidence-based Jaccard partition, but compositional constraints should be borne in mind when interpreting per-pathogen dispersion. Finally, absolute JSD values are small; the finding is a large relative change from a near-zero baseline, and its clinical significance scales with the absolute levels reported. Likewise, with five timepoints the joinpoint analysis could test at most one breakpoint; annual national series, when released, would allow finer temporal localisation of any acceleration.

## 5. Conclusions

National childhood respiratory pathogen spectra have moved apart as they have diversified. Between 1990 and 2023, mean pairwise compositional distance among countries roughly tripled (+238% in Jensen–Shannon divergence, +120% in Bray–Curtis dissimilarity), a result that held across distance metrics, pathogen panels, exclusion of COVID-19, low-count thresholds and a fixed balanced panel of 107 countries, where the increase was steepest (+423%). The divergence peaked during the 2021 pandemic window and receded only partially by 2023, suggesting a reversible shock component superimposed on a persistent secular trend. Decomposition shows that the divergence is reallocation of shares within a slowly expanding common pathogen set, not the appearance of country-specific subsets; it is fastest for viral pathogens and for pneumococcus itself; and it occurs within every assessable world region rather than between regional blocs.

Diversification without convergence carries a coherent implication for transition theory: at the etiologic-spectrum level, the childhood respiratory transition is asynchronous and path-dependent, and there is no single destination spectrum toward which countries are travelling. The shared pneumococcus-dominated starting point of 1990 has dissolved into increasingly country-specific compositions as vaccine-preventable dominance recedes at different speeds. For policy, the era in which one empirical spectrum could anchor treatment algorithms, surveillance priorities and vaccine strategies across countries is receding; compositional surveillance at national and regional scale becomes correspondingly more valuable, and shared guidelines should be treated as starting hypotheses to be locally verified rather than as descriptions of local reality. For monitoring, paired alpha- and beta-diversity reporting offers a compact, transferable framework for tracking how the structure of disease burden changes as it declines, applicable wherever multi-cause mortality or morbidity spectra are estimated.

## Data Availability

Derived national share panels and pairwise distance series are available from the corresponding author; all inputs from the GBD 2023 Results Tool.

https://vizhub.healthdata.org/gbd-results/

## Declarations

### Ethics approval

Not required; the study uses publicly available, de-identified, aggregate modelled estimates.

### Derived national share panels and pairwise distance series are available from the corresponding author; a

Funding This work was supported by the Beijing Science and Technology Nova Program Interdisciplinary Project (20230484439) and the Beijing High-Level Public Health Technical Talents Project (2023-02-33). The funder had no role in study design, data collection, data analysis, data interpretation, or writing of the report.

### Presentation

This work has not been presented at any scientific meeting.

Disclosure The authors declare no conflicts of interest. AI tools were used for language editing and for assistance with code generation and modification; the authors take full responsibility for the content.

### Patient and public involvement

Patients and the public were not involved in this secondary analysis of modelled estimates.

Contributors SC conceived the study, led the research design, and is the principal investigator responsible for the study idea and overall conduct. DL and HC curated the data and performed the analysis. SC drafted the manuscript. SC supervised the study and is the corresponding author. All authors revised the manuscript and approved the final version. SC is the guarantor.

## References

1. Omran AR. The epidemiologic transition: a theory of the epidemiology of population change. Milbank Mem Fund Q. 1971;49(4):509–538. doi:10.2307/3349375.

2. Mackenbach JP. Omran’s ‘Epidemiologic Transition’ 50 years on. Int J Epidemiol. 2022;51(3):1054–1057. doi:10.1093/ije/dyac020.

3. Santosa A, Wall S, Fottrell E, Högberg U, Byass P. The development and experience of epidemiological transition theory over four decades: a systematic review. Glob Health Action. 2014;7:23574. doi:10.3402/gha.v7.23574.

4. Jamison DT, Summers LH, Alleyne G, et al. Global health 2035: a world converging within a generation. Lancet. 2013;382(9908):1898–1955. doi:10.1016/S0140-6736(13)62105-4.

5. Vallin J, Meslé F. Convergences and divergences in mortality: a new approach to health transition. Demogr Res. 2004;Special Collection 2:11–44. doi:10.4054/DemRes.2004.S2.2.

6. Moser K, Shkolnikov V, Leon DA. World mortality 1950–2000: divergence replaces convergence from the late 1980s. Bull World Health Organ. 2005;83(3):202–209.

7. McMichael AJ, McKee M, Shkolnikov V, Valkonen T. Mortality trends and setbacks: global convergence or divergence? Lancet. 2004;363(9415):1155–1159. doi:10.1016/S0140-6736(04)15902-3.

8. Atance D, Claramunt MM, Varea X, Aburto JM. Convergence and divergence in mortality: a global study from 1990 to 2030. PLoS One. 2024;19(1):e0295842. doi:10.1371/journal.pone.0295842.

9. GBD 2023 Causes of Death Collaborators. Global burden of 292 causes of death in 204 countries and territories and 660 subnational locations, 1990–2023: a systematic analysis for the Global Burden of Disease Study 2023. Lancet. 2025;406(10513):1811–1872. doi:10.1016/S0140-6736(25)01917-8.

10. Wahl B, O’Brien KL, Greenbaum A, et al. Burden of Streptococcus pneumoniae and Haemophilus influenzae type b disease in children in the era of conjugate vaccines: global, regional, and national estimates for 2000–15. Lancet Glob Health. 2018;6(7):e744–e757. doi:10.1016/S2214-109X(18)30247-X.

11. Whittaker RH. Vegetation of the Siskiyou Mountains, Oregon and California. Ecol Monogr. 1960;30(3):279–338. doi:10.2307/1943563.

12. World Health Organization. Pocket book of hospital care for children: guidelines for the management of common childhood illnesses. 2nd ed. Geneva: World Health Organization; 2013.

13. GBD 2021 Lower Respiratory Infections and Antimicrobial Resistance Collaborators. Global, regional, and national incidence and mortality burden of non-COVID-19 lower respiratory infections and aetiologies, 1990–2021. Lancet Infect Dis. 2024;24(9):974–1002. doi:10.1016/S1473-3099(24)00176-2.

14. GBD 2023 Lower Respiratory Infections and Antimicrobial Resistance Collaborators. Global burden of lower respiratory infections and aetiologies, 1990–2023: a systematic analysis for the Global Burden of Disease Study 2023. Lancet Infect Dis. 2026;26(4):343–361. doi:10.1016/S1473-3099(25)00689-9.

15. Lin J. Divergence measures based on the Shannon entropy. IEEE Trans Inf Theory. 1991;37(1):145–151. doi:10.1109/18.61115.

16. Bray JR, Curtis JT. An ordination of the upland forest communities of southern Wisconsin. Ecol Monogr. 1957;27(4):325–349. doi:10.2307/1942268.

17. Baselga A. Separating the two components of abundance-based dissimilarity: balanced changes in abundance vs. abundance gradients. Methods Ecol Evol. 2013;4(6):552–557. doi:10.1111/2041-210X.12029.

18. Baselga A. Partitioning the turnover and nestedness components of beta diversity. Glob Ecol Biogeogr. 2010;19(1):134–143. doi:10.1111/j.1466-8238.2009.00490.x.

19. World Health Organization and UNICEF. WHO/UNICEF estimates of national immunization coverage (WUENIC), 2025 revision. Geneva: World Health Organization; 2026. Available at: https://immunizationdata.who.int (PCV3 and Hib3 coverage indicators).

20. Chow EJ, Uyeki TM, Chu HY. The effects of the COVID-19 pandemic on community respiratory virus activity. Nat Rev Microbiol. 2023;21(3):195–210. doi:10.1038/s41579-022-00807-9.

21. Bergeron-Boucher MP, Aburto JM, van Raalte A. Diversification in causes of death in low-mortality countries: emerging patterns and implications. BMJ Glob Health. 2020;5(7):e002414. doi:10.1136/bmjgh-2020-002414.

22. Calazans JA, Permanyer I. Levels, trends, and determinants of cause-of-death diversity in a global perspective: 1990–2019. BMC Public Health. 2023;23:650. doi:10.1186/s12889-023-15502-4.

23. GBD 2023 Disease and Injury and Risk Factor Collaborators. Burden of 375 diseases and injuries, risk-attributable burden of 88 risk factors, and healthy life expectancy in 204 countries and territories and 660 subnational locations, 1990–2023: a systematic analysis for the Global Burden of Disease Study 2023. Lancet. 2025;406(10513):1873–1922. doi:10.1016/S0140-6736(25)01637-X.

