## Supplementary_Materials for "Diversification without convergence: national childhood respiratory pathogen spectra diverge as they diversify, 1990–2023"

Contents: Supplementary Tables S1–S6 and Supplementary Note S7 (panel methods). All estimates derive from the Global Burden of Disease Study 2023 (GBD 2023, release v8352), ages 0–19 years, both sexes combined.

**Supplementary Table S1. Pairwise between-country compositional distance by timepoint and metric: full summary table (26-pathogen panel, 100-death low-count filter).**

| Year | Countries, n | JSD mean | JSD median | JSD Q1 | JSD Q3 | Bray–Curtis mean | Bray–Curtis median | Bray–Curtis Q1 | Bray–Curtis Q3 |
|---|---|---|---|---|---|---|---|---|---|
| 1990 | 137 | 0.0084 | 0.0051 | 0.0020 | 0.0110 | 0.066 | 0.061 | 0.040 | 0.087 |
| 2010 | 115 | 0.0136 | 0.0083 | 0.0035 | 0.0162 | 0.090 | 0.080 | 0.052 | 0.112 |
| 2019 | 111 | 0.0250 | 0.0183 | 0.0084 | 0.0343 | 0.139 | 0.130 | 0.086 | 0.183 |
| 2021 | 107 | 0.0320 | 0.0270 | 0.0146 | 0.0446 | 0.145 | 0.140 | 0.099 | 0.183 |
| 2023 | 108 | 0.0283 | 0.0240 | 0.0126 | 0.0384 | 0.145 | 0.138 | 0.100 | 0.182 |

JSD, Jensen–Shannon divergence (base 2, bounded [0, 1]); Q1/Q3, first and third quartiles of the $n(n − 1)/2$ pairwise values. Change 1990 to 2023: JSD mean +238% (linear trend $p = 0.030$); Bray–Curtis mean +120% ($p = 0.014$).

**Supplementary Table S2. Robustness matrix for the divergence trend (mean pairwise JSD): balanced panel (a), pathogen-set and COVID-19 calibres (b), and low-count thresholds (c).**

(a) Balanced panel: 107 countries passing the 100-death filter at all five timepoints.

| Year | Countries, n | JSD mean | JSD median |
|---|---|---|---|
| 1990 | 107 | 0.0052 | 0.0037 |
| 2010 | 107 | 0.0109 | 0.0074 |
| 2019 | 107 | 0.0219 | 0.0170 |
| 2021 | 107 | 0.0320 | 0.0270 |
| 2023 | 107 | 0.0275 | 0.0235 |

(b) Pathogen-set sensitivity: 26-pathogen panel versus 29-pathogen panel (adding tuberculosis, pertussis and COVID-19), and the 29-pathogen panel excluding COVID-19.

| Year | n (26) | JSD mean (26) | JSD median (26) | n (29) | JSD mean (29) | JSD median (29) | n (29 excl. COVID - 19) | JSD mean (29 excl. COVID - 19) | JSD median (29 excl. COVID - 19) |
|---|---|---|---|---|---|---|---|---|---|
| 2010 | 115 | 0.0136 | 0.0083 | 118 | 0.056 | 0.042 | 118 | 0.056 | 0.042 |
| 2019 | 111 | 0.0250 | 0.0183 | 111 | 0.067 | 0.053 | 111 | 0.067 | 0.053 |
| 2021 | 107 | 0.0320 | 0.0270 | 112 | 0.103 | 0.086 | 112 | 0.072 | 0.061 |

(c) Low-count threshold sensitivity: minimum total attributed deaths required for inclusion.

| Threshold (deaths) | Year | Countries, n | JSD mean | JSD median |
|---|---|---|---|---|
| 50 | 1990 | 152 | 0.0092 | 0.0056 |

| Threshold (deaths) | Year | Countries, n | JSD mean | JSD median |
|---|---|---|---|---|
| 50 | 2010 | 133 | 0.0194 | 0.0107 |
| 50 | 2019 | 127 | 0.0291 | 0.0207 |
| 50 | 2021 | 120 | 0.0368 | 0.0303 |
| 50 | 2023 | 121 | 0.0327 | 0.0266 |
| 100 | 1990 | 137 | 0.0084 | 0.0051 |
| 100 | 2010 | 115 | 0.0136 | 0.0083 |
| 100 | 2019 | 111 | 0.0250 | 0.0183 |
| 100 | 2021 | 107 | 0.0320 | 0.0270 |
| 100 | 2023 | 108 | 0.0283 | 0.0240 |
| 200 | 1990 | 123 | 0.0078 | 0.0048 |
| 200 | 2010 | 105 | 0.0114 | 0.0074 |
| 200 | 2019 | 101 | 0.0215 | 0.0163 |
| 200 | 2021 | 93 | 0.0312 | 0.0261 |
| 200 | 2023 | 96 | 0.0266 | 0.0221 |

Change 1990 to 2023 in mean pairwise JSD: +423% (balanced panel, trend $p = 0.038$); +255%, +238% and +243% under thresholds 50, 100 and 200 deaths, respectively. Pertussis country estimates are unavailable for 1990 and 2023, so the 29-pathogen calibres span 2010–2021 only; COVID-19 is zero before 2020 by construction. For 2010 and 2019 the 29-pathogen panel excluding COVID-19 is identical to the full 29-pathogen panel.

### Supplementary Table S3. Beta-diversity decomposition of between-country dissimilarity:

Baselga abundance partition of Bray–Curtis dissimilarity (a) and Jaccard partition into turnover and nestedness components (b), 1990–2023.

(a) Baselga abundance partition (Bray–Curtis).

| Year | Bray–Curtis dissimilarity | Balanced variation component | Gradient component | Balanced variation, % | Gradient, % |
|---|---|---|---|---|---|
| 1990 | 0.0657 | 0.0657 | 0 (identity) | 100 | 0 |
| 2010 | 0.0900 | 0.0900 | 0 (identity) | 100 | 0 |
| 2019 | 0.1393 | 0.1393 | 0 (identity) | 100 | 0 |
| 2021 | 0.1451 | 0.1451 | 0 (identity) | 100 | 0 |
| 2023 | 0.1446 | 0.1446 | 0 (identity) | 100 | 0 |

(b) Jaccard partition (presence defined as share ≥0.5%).

| Year | Jaccard dissimilarity | Turnover component | Nestedness component | Turnover fraction, % | Mean richness (of 26) |
|---|---|---|---|---|---|
| 1990 | 0.052 | 0.022 | 0.030 | 42.8 | 18.5 |
| 2010 | 0.061 | 0.007 | 0.054 | 11.2 | 19.2 |
| 2019 | 0.052 | 0.022 | 0.030 | 42.2 | 20.7 |
| 2021 | 0.089 | 0.029 | 0.060 | 32.8 | 20.9 |
| 2023 | 0.059 | 0.019 | 0.040 | 31.8 | 21.1 |

For closed share vectors both abundance totals equal one, so the gradient component is identically zero (residual floating-point values $<10^{-16}$) and the 100%/0% split is an identity by construction, reported for completeness rather than as an empirical result. The nestedness fraction is the complement of the turnover fraction.

**Supplementary Table S4. Cross-country dispersion of national pathogen shares, full 26- pathogen panel by timepoint: number of included countries, mean share, coefficient of variation (CV) and interquartile range (IQR) of share.**

| Year | Pathogen | Countries, n | Mean share, % | CV | IQR of share, percentage points |
|---|---|---|---|---|---|
| 1990 | Acinetobacter baumannii | 137 | 1.78 | 0.24 | 0.67 |
| 1990 | Aspergillus spp. | 137 | 1.38 | 0.39 | 0.52 |
| 1990 | Chlamydia spp | 137 | 1.47 | 0.39 | 0.91 |
| 1990 | Citrobacter spp. | 137 | 0.14 | 0.45 | 0.07 |
| 1990 | Enterobacter spp | 137 | 0.40 | 0.16 | 0.09 |
| 1990 | Escherichia coli | 137 | 3.21 | 0.11 | 0.46 |
| 1990 | Group A Streptococcus | 137 | 2.67 | 0.33 | 1.33 |
| 1990 | Group B streptococcus | 137 | 0.85 | 0.27 | 0.34 |
| 1990 | Haemophilus influenzae | 137 | 2.25 | 0.16 | 0.52 |
| 1990 | Influenza | 137 | 4.58 | 0.03 | 0.17 |
| 1990 | Klebsiella pneumoniae | 137 | 8.89 | 0.19 | 2.29 |
| 1990 | Legionella spp | 137 | 0.11 | 2.86 | 0.04 |
| 1990 | Morganella spp. | 137 | 0.09 | 0.13 | 0.02 |
| 1990 | Mycoplasma | 137 | 2.76 | 0.07 | 0.19 |
| 1990 | Other Acinetobacter species | 137 | 0.41 | 0.04 | 0.02 |
| 1990 | Other Klebsiella species | 137 | 0.47 | 0.21 | 0.18 |
| 1990 | Other Mycobacterium species (non- TB, non- Leprosy) | 137 | 2.81 | 0.17 | 0.59 |
| 1990 | Other bacterial and viral pathogens | 137 | 1.42 | 0.25 | 0.50 |
| 1990 | Other fungi | 137 | 0.87 | 0.29 | 0.21 |
| 1990 | Other gram-negative bacteria | 137 | 1.83 | 0.06 | 0.21 |
| 1990 | Proteus spp. | 137 | 0.10 | 0.26 | 0.02 |
| 1990 | Pseudomonas aeruginosa | 137 | 4.84 | 0.07 | 0.44 |
| 1990 | Respiratory syncytial virus | 137 | 4.51 | 0.08 | 0.46 |
| 1990 | Serratia spp. | 137 | 0.20 | 0.13 | 0.04 |
| 1990 | Staphylococcus aureus | 137 | 3.34 | 0.32 | 1.25 |
| 1990 | Streptococcus pneumoniae | 137 | 48.62 | 0.06 | 4.02 |

| Year | Pathogen | Countries, n | Mean share, % | CV | IQR of share, percentage points |
|---|---|---|---|---|---|
| 2010 | Acinetobacter baumannii | 115 | 1.67 | 0.25 | 0.68 |
| 2010 | Aspergillus spp. | 115 | 1.70 | 0.37 | 0.80 |
| 2010 | Chlamydia spp | 115 | 1.38 | 0.37 | 0.83 |
| 2010 | Citrobacter spp. | 115 | 0.20 | 0.50 | 0.13 |
| 2010 | Enterobacter spp | 115 | 0.50 | 0.20 | 0.11 |
| 2010 | Escherichia coli | 115 | 3.87 | 0.16 | 0.81 |
| 2010 | Group A Streptococcus | 115 | 2.74 | 0.32 | 1.33 |
| 2010 | Group B streptococcus | 115 | 0.98 | 0.24 | 0.31 |
| 2010 | Haemophilus influenzae | 115 | 2.13 | 0.20 | 0.60 |
| 2010 | Influenza | 115 | 4.73 | 0.12 | 0.21 |
| 2010 | Klebsiella pneumoniae | 115 | 9.16 | 0.17 | 2.28 |
| 2010 | Legionella spp | 115 | 0.22 | 2.79 | 0.16 |
| 2010 | Morganella spp. | 115 | 0.10 | 0.14 | 0.02 |
| 2010 | Mycoplasma | 115 | 2.89 | 0.09 | 0.22 |
| 2010 | Other Acinetobacter species | 115 | 0.43 | 0.07 | 0.04 |
| 2010 | Other Klebsiella species | 115 | 0.60 | 0.25 | 0.22 |
| 2010 | Other Mycobacterium species (non- TB, non- Leprosy) | 115 | 3.42 | 0.18 | 0.71 |
| 2010 | Other bacterial and viral pathogens | 115 | 1.60 | 0.21 | 0.48 |
| 2010 | Other fungi | 115 | 0.91 | 0.28 | 0.27 |
| 2010 | Other gram-negative bacteria | 115 | 2.03 | 0.12 | 0.30 |
| 2010 | Proteus spp. | 115 | 0.14 | 0.38 | 0.06 |
| 2010 | Pseudomonas aeruginosa | 115 | 5.35 | 0.09 | 0.53 |
| 2010 | Respiratory syncytial virus | 115 | 4.99 | 0.13 | 0.76 |
| 2010 | Serratia spp. | 115 | 0.27 | 0.25 | 0.08 |
| 2010 | Staphylococcus aureus | 115 | 4.72 | 0.43 | 2.45 |
| 2010 | Streptococcus pneumoniae | 115 | 43.26 | 0.13 | 5.45 |
| 2019 | Acinetobacter baumannii | 111 | 1.38 | 0.29 | 0.57 |
| 2019 | Aspergillus spp. | 111 | 1.90 | 0.33 | 0.82 |
| 2019 | Chlamydia spp | 111 | 1.58 | 0.39 | 1.04 |
| 2019 | Citrobacter spp. | 111 | 0.26 | 0.44 | 0.18 |
| 2019 | Enterobacter spp | 111 | 0.63 | 0.16 | 0.14 |
| 2019 | Escherichia coli | 111 | 4.88 | 0.14 | 0.87 |
| 2019 | Group A Streptococcus | 111 | 2.99 | 0.29 | 1.22 |

| Year | Pathogen | Countries, n | Mean share, % | CV | IQR of share, percentage points |
|---|---|---|---|---|---|
| 2019 | Group B streptococcus | 111 | 1.30 | 0.25 | 0.46 |
| 2019 | Haemophilus influenzae | 111 | 2.31 | 0.17 | 0.47 |
| 2019 | Influenza | 111 | 7.43 | 0.37 | 4.29 |
| 2019 | Klebsiella pneumoniae | 111 | 10.08 | 0.18 | 2.78 |
| 2019 | Legionella spp | 111 | 0.33 | 2.34 | 0.32 |
| 2019 | Morganella spp. | 111 | 0.12 | 0.13 | 0.02 |
| 2019 | Mycoplasma | 111 | 3.59 | 0.14 | 0.79 |
| 2019 | Other Acinetobacter species | 111 | 0.54 | 0.17 | 0.17 |
| 2019 | Other Klebsiella species | 111 | 0.82 | 0.20 | 0.22 |
| 2019 | Other Mycobacterium species (non- TB, non- Leprosy) | 111 | 4.47 | 0.15 | 0.92 |
| 2019 | Other bacterial and viral pathogens | 111 | 1.89 | 0.19 | 0.49 |
| 2019 | Other fungi | 111 | 1.10 | 0.28 | 0.32 |
| 2019 | Other gram-negative bacteria | 111 | 2.53 | 0.17 | 0.65 |
| 2019 | Proteus spp. | 111 | 0.20 | 0.29 | 0.08 |
| 2019 | Pseudomonas aeruginosa | 111 | 6.65 | 0.12 | 1.34 |
| 2019 | Respiratory syncytial virus | 111 | 7.14 | 0.26 | 2.70 |
| 2019 | Serratia spp. | 111 | 0.38 | 0.18 | 0.10 |
| 2019 | Staphylococcus aureus | 111 | 6.23 | 0.33 | 3.26 |
| 2019 | Streptococcus pneumoniae | 111 | 29.26 | 0.28 | 12.55 |
| 2021 | Acinetobacter baumannii | 107 | 1.51 | 0.24 | 0.49 |
| 2021 | Aspergillus spp. | 107 | 2.20 | 0.34 | 1.05 |
| 2021 | Chlamydia spp | 107 | 1.81 | 0.36 | 1.05 |
| 2021 | Citrobacter spp. | 107 | 0.30 | 0.44 | 0.22 |
| 2021 | Enterobacter spp | 107 | 0.72 | 0.18 | 0.17 |
| 2021 | Escherichia coli | 107 | 5.58 | 0.16 | 1.13 |
| 2021 | Group A Streptococcus | 107 | 3.35 | 0.30 | 1.51 |
| 2021 | Group B streptococcus | 107 | 1.51 | 0.24 | 0.52 |
| 2021 | Haemophilus influenzae | 107 | 2.59 | 0.21 | 0.60 |
| 2021 | Influenza | 107 | 2.66 | 0.83 | 3.42 |
| 2021 | Klebsiella pneumoniae | 107 | 11.33 | 0.16 | 2.50 |
| 2021 | Legionella spp | 107 | 0.30 | 1.84 | 0.37 |
| 2021 | Morganella spp. | 107 | 0.14 | 0.12 | 0.02 |

| Year | Pathogen | Countries, n | Mean share, % | CV | IQR of share, percentage points |
|---|---|---|---|---|---|
| 2021 | Mycoplasma | 107 | 4.14 | 0.17 | 0.96 |
| 2021 | Other Acinetobacter species | 107 | 0.63 | 0.20 | 0.21 |
| 2021 | Other Klebsiella species | 107 | 0.95 | 0.22 | 0.27 |
| 2021 | Other Mycobacterium species (non- TB, non- Leprosy) | 107 | 5.32 | 0.16 | 1.22 |
| 2021 | Other bacterial and viral pathogens | 107 | 2.20 | 0.18 | 0.59 |
| 2021 | Other fungi | 107 | 1.32 | 0.30 | 0.46 |
| 2021 | Other gram-negative bacteria | 107 | 2.86 | 0.17 | 0.70 |
| 2021 | Proteus spp. | 107 | 0.25 | 0.31 | 0.09 |
| 2021 | Pseudomonas aeruginosa | 107 | 7.64 | 0.15 | 1.81 |
| 2021 | Respiratory syncytial virus | 107 | 2.27 | 0.75 | 2.77 |
| 2021 | Serratia spp. | 107 | 0.44 | 0.20 | 0.12 |
| 2021 | Staphylococcus aureus | 107 | 7.21 | 0.32 | 3.56 |
| 2021 | Streptococcus pneumoniae | 107 | 30.79 | 0.25 | 11.23 |
| 2023 | Acinetobacter baumannii | 108 | 1.45 | 0.27 | 0.47 |
| 2023 | Aspergillus spp. | 108 | 2.10 | 0.33 | 0.99 |
| 2023 | Chlamydia spp | 108 | 1.76 | 0.39 | 1.07 |
| 2023 | Citrobacter spp. | 108 | 0.29 | 0.43 | 0.19 |
| 2023 | Enterobacter spp | 108 | 0.70 | 0.16 | 0.16 |
| 2023 | Escherichia coli | 108 | 5.42 | 0.15 | 0.96 |
| 2023 | Group A Streptococcus | 108 | 3.26 | 0.29 | 1.34 |
| 2023 | Group B streptococcus | 108 | 1.47 | 0.26 | 0.52 |
| 2023 | Haemophilus influenzae | 108 | 2.52 | 0.18 | 0.55 |
| 2023 | Influenza | 108 | 4.34 | 0.55 | 3.33 |
| 2023 | Klebsiella pneumoniae | 108 | 11.01 | 0.18 | 2.82 |
| 2023 | Legionella spp | 108 | 0.32 | 2.04 | 0.37 |
| 2023 | Morganella spp. | 108 | 0.13 | 0.14 | 0.03 |
| 2023 | Mycoplasma | 108 | 3.98 | 0.17 | 0.98 |
| 2023 | Other Acinetobacter species | 108 | 0.61 | 0.21 | 0.21 |
| 2023 | Other Klebsiella species | 108 | 0.91 | 0.21 | 0.25 |
| 2023 | Other Mycobacterium species (non- TB, non- Leprosy) | 108 | 5.12 | 0.16 | 0.98 |

| Year | Pathogen | Countries, n | Mean share, % | CV | IQR of share, percentage points |
|---|---|---|---|---|---|
| 2023 | Other bacterial and viral pathogens | 108 | 2.13 | 0.19 | 0.57 |
| 2023 | Other fungi | 108 | 1.26 | 0.29 | 0.42 |
| 2023 | Other gram-negative bacteria | 108 | 2.76 | 0.19 | 0.73 |
| 2023 | Proteus spp. | 108 | 0.24 | 0.30 | 0.08 |
| 2023 | Pseudomonas aeruginosa | 108 | 7.40 | 0.15 | 1.90 |
| 2023 | Respiratory syncytial virus | 108 | 3.97 | 0.48 | 2.59 |
| 2023 | Serratia spp. | 108 | 0.43 | 0.18 | 0.11 |
| 2023 | Staphylococcus aureus | 108 | 6.96 | 0.31 | 3.34 |
| 2023 | Streptococcus pneumoniae | 108 | 29.47 | 0.26 | 10.42 |

CV, coefficient of variation (standard deviation divided by mean) across included countries; IQR, interquartile range. 15 of 26 pathogens showed rising CV between 1990 and 2023. Shares are proportions of the country-timepoint total of the 26 rei-level PAF-attributed LRI etiologies.

**Supplementary Table S5. Regional stratification: mean pairwise JSD within each GBD super- region (a) and between country pairs spanning different super-regions (b), 1990–2023. (a) Within-super-region pairwise JSD.**

| Year | Super-region | Countries, n | Within-region JSD mean | Within-region JSD median |
|---|---|---|---|---|
| 1990 | Central Europe, Eastern Europe, and Central Asia | 23 | 0.0045 | 0.0033 |
| 1990 | High-income | 12 | 0.0064 | 0.0047 |
| 1990 | Latin America and Caribbean | 19 | 0.0041 | 0.0025 |
| 1990 | North Africa and Middle East | 18 | 0.0035 | 0.0023 |
| 1990 | South Asia | 5 | 0.0009 | 0.0007 |
| 1990 | Southeast Asia, East Asia, and Oceania | 16 | 0.0052 | 0.0039 |
| 1990 | Sub-Saharan Africa | 44 | 0.0021 | 0.0012 |
| 2010 | Central Europe, Eastern Europe, and Central Asia | 15 | 0.0047 | 0.0038 |
| 2010 | High-income | 5 | 0.0337 | 0.0354 |
| 2010 | Latin America and Caribbean | 16 | 0.0067 | 0.0045 |
| 2010 | North Africa and Middle East | 16 | 0.0089 | 0.0072 |
| 2010 | South Asia | 5 | 0.0020 | 0.0018 |

| Year | Super-region | Countries, n | Within-region JSD mean | Within-region JSD median |
|---|---|---|---|---|
| 2010 | Southeast Asia, East Asia, and Oceania | 14 | 0.0172 | 0.0088 |
| 2010 | Sub-Saharan Africa | 44 | 0.0044 | 0.0024 |
| 2019 | Central Europe, Eastern Europe, and Central Asia | 12 | 0.0183 | 0.0169 |
| 2019 | Latin America and Caribbean | 16 | 0.0165 | 0.0105 |
| 2019 | North Africa and Middle East | 17 | 0.0242 | 0.0186 |
| 2019 | Southeast Asia, East Asia, and Oceania | 14 | 0.0274 | 0.0147 |
| 2019 | Sub-Saharan Africa | 44 | 0.0130 | 0.0078 |
| 2021 | Central Europe, Eastern Europe, and Central Asia | 12 | 0.0243 | 0.0213 |
| 2021 | Latin America and Caribbean | 16 | 0.0264 | 0.0192 |
| 2021 | North Africa and Middle East | 16 | 0.0196 | 0.0179 |
| 2021 | Southeast Asia, East Asia, and Oceania | 13 | 0.0181 | 0.0161 |
| 2021 | Sub-Saharan Africa | 44 | 0.0171 | 0.0135 |
| 2023 | Central Europe, Eastern Europe, and Central Asia | 12 | 0.0197 | 0.0162 |
| 2023 | Latin America and Caribbean | 16 | 0.0231 | 0.0193 |
| 2023 | North Africa and Middle East | 16 | 0.0210 | 0.0178 |
| 2023 | Southeast Asia, East Asia, and Oceania | 13 | 0.0182 | 0.0148 |
| 2023 | Sub-Saharan Africa | 44 | 0.0190 | 0.0145 |

(b) Between-super-region pairwise JSD (country pairs in different super-regions).

| Year | Between-region JSD mean | Between-region JSD median |
|---|---|---|
| 1990 | 0.0096 | 0.0062 |
| 2010 | 0.0157 | 0.0101 |
| 2019 | 0.0277 | 0.0212 |
| 2021 | 0.0362 | 0.0317 |
| 2023 | 0.0310 | 0.0260 |

Within-region statistics are computed only where at least five included countries remain in the super-region; n = 29 super-region-by-timepoint cells meet this criterion. Fold changes 1990 to 2023 for the five continuously computable super-regions range from ×3.5 (Southeast Asia, East Asia and Oceania) to ×9.0 (Sub-Saharan Africa); between-region pairs rose ×3.2.

**Supplementary Table S6. Low-count filter and country lists: exclusions under the 100-death threshold and the countries included at each timepoint (26-pathogen panel).**

| Panel / timepoint | 1990 | 2010 | 2019 | 2021 | 2023 |
|---|---|---|---|---|---|
| 26-pathogen panel: countries excluded (<100 deaths) | 67 | 89 | 93 | 97 | 96 |
| 26-pathogen panel: countries included | 137 | 115 | 111 | 107 | 108 |
| 29-pathogen panel: countries excluded (<100 deaths) | n/a | 86 | 93 | 92 | n/a |
| 29-pathogen panel: countries included | n/a | 118 | 111 | 112 | n/a |

n/a, not applicable (pertussis country estimates unavailable for 1990 and 2023). Exclusions rise over time as mortality decline pushes predominantly high-income and small states below the filter.

Included countries, 1990 (n = 137). Afghanistan, Albania, Algeria, Angola, Argentina, Armenia, Azerbaijan, Bangladesh, Belarus, Benin, Bhutan, Bolivia (Plurinational State of), Bosnia and Herzegovina, Botswana, Brazil, Bulgaria, Burkina Faso, Burundi, Cambodia, Cameroon, Canada, Central African Republic, Chad, Chile, China, Colombia, Comoros, Congo, Costa Rica, Cuba, Czechia, Côte d'Ivoire, Democratic People's Republic of Korea, Democratic Republic of the Congo, Djibouti, Dominican Republic, Ecuador, Egypt, El Salvador, Equatorial Guinea, Eritrea, Eswatini, Ethiopia, Fiji, France, Gabon, Gambia, Georgia, Germany, Ghana, Guatemala, Guinea, Guinea- Bissau, Guyana, Haiti, Honduras, Hungary, India, Indonesia, Iran (Islamic Republic of), Iraq, Italy, Jamaica, Japan, Jordan, Kazakhstan, Kenya, Kyrgyzstan, Lao People's Democratic Republic, Lebanon, Lesotho, Liberia, Libya, Madagascar, Malawi, Malaysia, Maldives, Mali, Mauritania, Mexico, Mongolia, Morocco, Mozambique, Myanmar, Namibia, Nepal, Nicaragua, Niger, Nigeria, North Macedonia, Oman, Pakistan, Palestine, Panama, Papua New Guinea, Paraguay, Peru, Philippines, Poland, Portugal, Republic of Korea, Republic of Moldova, Romania, Russian Federation, Rwanda, Saudi Arabia, Senegal, Serbia, Sierra Leone, Slovakia, Somalia, South Africa, South Sudan, Spain, Sri Lanka, Sudan, Syrian Arab Republic, Taiwan, Tajikistan, Thailand, Timor-Leste, Togo, Tunisia, Turkmenistan, Türkiye, Uganda, Ukraine, United Arab Emirates, United Kingdom, United Republic of Tanzania, United States of America, Uzbekistan, Venezuela (Bolivarian Republic of), Viet Nam, Yemen, Zambia, Zimbabwe.

Included countries, 2010 (n = 115). Afghanistan, Algeria, Angola, Argentina, Armenia, Azerbaijan, Bangladesh, Benin, Bhutan, Bolivia (Plurinational State of), Botswana, Brazil, Bulgaria, Burkina Faso, Burundi, Cambodia, Cameroon, Central African Republic, Chad, Chile, China, Colombia, Comoros, Congo, Cuba, Côte d'Ivoire, Democratic People's Republic of Korea, Democratic Republic of the Congo, Djibouti, Dominican Republic, Ecuador, Egypt, El Salvador, Equatorial Guinea, Eritrea, Eswatini, Ethiopia, Gabon, Gambia, Georgia, Ghana, Guatemala, Guinea, Guinea-Bissau, Haiti, Honduras, India, Indonesia, Iran (Islamic Republic of), Iraq, Japan, Jordan, Kazakhstan, Kenya, Kyrgyzstan, Lao People's Democratic Republic, Lebanon, Lesotho, Liberia, Libya, Madagascar, Malawi, Malaysia, Mali, Mauritania, Mexico, Mongolia, Morocco, Mozambique, Myanmar, Namibia, Nepal, Nicaragua, Niger, Nigeria, Pakistan, Palestine, Panama, Papua New Guinea, Paraguay, Peru, Philippines, Poland, Republic of Moldova, Romania, Russian Federation, Rwanda, Saudi Arabia, Senegal, Sierra Leone, Somalia, South Africa, South Sudan, Sri Lanka, Sudan, Syrian Arab Republic, Taiwan, Tajikistan, Thailand, Timor-Leste, Togo, Tunisia, Turkmenistan, Türkiye, Uganda, Ukraine, United Kingdom, United Republic of Tanzania, United States of America, Uzbekistan, Venezuela (Bolivarian Republic of), Viet Nam, Yemen, Zambia, Zimbabwe.

Included countries, 2019 (n = 111). Afghanistan, Algeria, Angola, Argentina, Armenia, Azerbaijan, Bangladesh, Benin, Bolivia (Plurinational State of), Botswana, Brazil, Burkina Faso, Burundi, Cambodia, Cameroon, Central African Republic, Chad, China, Colombia, Comoros, Congo, Cuba, Côte d'Ivoire, Democratic People's Republic of Korea, Democratic Republic of the Congo, Djibouti, Dominican Republic, Ecuador, Egypt, El Salvador, Equatorial Guinea, Eritrea, Eswatini, Ethiopia, Gabon, Gambia, Ghana, Guatemala, Guinea, Guinea-Bissau, Haiti, Honduras, India, Indonesia, Iran (Islamic Republic of), Iraq, Japan, Jordan, Kazakhstan, Kenya, Kyrgyzstan, Lao People's Democratic Republic, Lebanon, Lesotho, Liberia, Libya, Madagascar, Malawi, Malaysia, Mali, Mauritania, Mexico, Mongolia, Morocco, Mozambique, Myanmar, Namibia, Nepal, Nicaragua, Niger, Nigeria, Pakistan, Palestine, Panama, Papua New Guinea, Paraguay, Peru, Philippines, Poland, Romania, Russian Federation, Rwanda, Saudi Arabia, Senegal, Sierra Leone, Somalia, South Africa, South Sudan, Sri Lanka, Sudan, Syrian Arab Republic, Taiwan, Tajikistan, Thailand, Timor-Leste, Togo, Tunisia, Turkmenistan, Türkiye, Uganda, Ukraine, United Arab Emirates, United Kingdom, United Republic of Tanzania, United States of America, Uzbekistan, Venezuela (Bolivarian Republic of), Viet Nam, Yemen, Zambia, Zimbabwe.

Included countries, 2021 (n = 107). Afghanistan, Algeria, Angola, Argentina, Armenia, Azerbaijan, Bangladesh, Benin, Bolivia (Plurinational State of), Botswana, Brazil, Burkina Faso, Burundi, Cambodia, Cameroon, Central African Republic, Chad, China, Colombia, Comoros, Congo, Cuba, Côte d'Ivoire, Democratic People's Republic of Korea, Democratic Republic of the Congo, Djibouti, Dominican Republic, Ecuador, Egypt, El Salvador, Equatorial Guinea, Eritrea, Eswatini, Ethiopia, Gabon, Gambia, Ghana, Guatemala, Guinea, Guinea-Bissau, Haiti, Honduras, India, Indonesia, Iran (Islamic Republic of), Iraq, Jordan, Kazakhstan, Kenya, Kyrgyzstan, Lao People's Democratic Republic, Lebanon, Lesotho, Liberia, Libya, Madagascar, Malawi, Malaysia, Mali, Mauritania, Mexico, Mongolia, Morocco, Mozambique, Myanmar, Namibia, Nepal, Nicaragua, Niger, Nigeria, Pakistan, Palestine, Panama, Papua New Guinea, Paraguay, Peru, Philippines, Poland, Romania, Russian Federation, Rwanda, Saudi Arabia, Senegal, Sierra Leone, Somalia, South Africa, South Sudan, Sri Lanka, Sudan, Syrian Arab Republic, Tajikistan, Thailand, Timor-Leste, Togo, Tunisia, Turkmenistan, Türkiye, Uganda, Ukraine, United Republic of Tanzania, United States of America, Uzbekistan, Venezuela (Bolivarian Republic of), Viet Nam, Yemen, Zambia, Zimbabwe.

Included countries, 2023 (n = 108). Afghanistan, Algeria, Angola, Argentina, Armenia, Azerbaijan, Bangladesh, Benin, Bolivia (Plurinational State of), Botswana, Brazil, Burkina Faso, Burundi, Cambodia, Cameroon, Central African Republic, Chad, China, Colombia, Comoros, Congo, Cuba, Côte d'Ivoire, Democratic People's Republic of Korea, Democratic Republic of the Congo, Djibouti, Dominican Republic, Ecuador, Egypt, El Salvador, Equatorial Guinea, Eritrea, Eswatini, Ethiopia, Gabon, Gambia, Ghana, Guatemala, Guinea, Guinea-Bissau, Haiti, Honduras, India, Indonesia, Iran (Islamic Republic of), Iraq, Jordan, Kazakhstan, Kenya, Kyrgyzstan, Lao People's Democratic Republic, Lebanon, Lesotho, Liberia, Libya, Madagascar, Malawi, Malaysia, Mali, Mauritania, Mexico, Mongolia, Morocco, Mozambique, Myanmar, Namibia, Nepal, Nicaragua, Niger, Nigeria, Pakistan, Palestine, Panama, Papua New Guinea, Paraguay, Peru, Philippines, Poland, Romania, Russian Federation, Rwanda, Saudi Arabia, Senegal, Sierra Leone, Somalia, South Africa, South Sudan, Sri Lanka, Sudan, Syrian Arab Republic, Tajikistan, Thailand, Timor-Leste, Togo, Tunisia, Turkmenistan, Türkiye, Uganda, Ukraine, United Kingdom, United Republic of Tanzania, United States of America, Uzbekistan, Venezuela (Bolivarian Republic of), Viet Nam, Yemen, Zambia, Zimbabwe.

Balanced panel (countries passing the 100-death filter at all five timepoints, n = 107). Afghanistan, Algeria, Angola, Argentina, Armenia, Azerbaijan, Bangladesh, Benin, Bolivia (Plurinational State of), Botswana, Brazil, Burkina Faso, Burundi, Cambodia, Cameroon, Central African Republic, Chad, China, Colombia, Comoros, Congo, Cuba, Côte d'Ivoire, Democratic People's Republic of Korea, Democratic Republic of the Congo, Djibouti, Dominican Republic, Ecuador, Egypt, El Salvador, Equatorial Guinea, Eritrea, Eswatini, Ethiopia, Gabon, Gambia, Ghana, Guatemala, Guinea, Guinea-Bissau, Haiti, Honduras, India, Indonesia, Iran (Islamic Republic of), Iraq, Jordan, Kazakhstan, Kenya, Kyrgyzstan, Lao People's Democratic Republic, Lebanon, Lesotho, Liberia, Libya, Madagascar, Malawi, Malaysia, Mali, Mauritania, Mexico, Mongolia, Morocco, Mozambique, Myanmar, Namibia, Nepal, Nicaragua, Niger, Nigeria, Pakistan, Palestine, Panama, Papua New Guinea, Paraguay, Peru, Philippines, Poland, Romania, Russian Federation, Rwanda, Saudi Arabia, Senegal, Sierra Leone, Somalia, South Africa, South Sudan, Sri Lanka, Sudan, Syrian Arab Republic, Tajikistan, Thailand, Timor-Leste, Togo, Tunisia, Turkmenistan, Türkiye, Uganda, Ukraine, United Republic of Tanzania, United States of America, Uzbekistan, Venezuela (Bolivarian Republic of), Viet Nam, Yemen, Zambia, Zimbabwe.

### Supplementary Note S7. Panel construction and analysis methods.

Data extraction. National pathogen-attributed lower respiratory infection (LRI) deaths were extracted from GBD 2023 (release v8352) at the risk–effect (rei) level for the 26 single-pathogen and residual etiologies, both sexes combined, for the four paediatric age groups (<5, 5–9, 10–14, 15–19 years) and five timepoints (1990, 2010, 2019, 2021, 2023). The rei-level interface returns central-estimate death counts from the counterfactual population-attributable-fraction attribution; uncertainty draws are not available at this interface. The four age groups were summed to a single 0–19 year total. For the 29-pathogen sensitivity panel, cause-level estimates of tuberculosis and pertussis (modelled directly by GBD as causes of death) and COVID-19 (2021 and 2023; zero before 2020 by construction) were concatenated; because GBD causes of death are mutually exclusive and collectively exhaustive, this introduces no double counting.

Spectrum vectors. For each country and timepoint, attributed deaths were divided by the country-timepoint total to form a closed compositional share vector over the pathogen panel. Country-timepoints with fewer than 100 total attributed deaths were excluded (primary filter; thresholds of 50 and 200 examined in sensitivity analysis). A balanced panel was defined as the 107 countries passing the 100-death filter at all five timepoints.

Distance metrics. For each timepoint, all pairwise Jensen–Shannon divergences (base-2 logarithms, bounded [0, 1]) among included countries were computed as $JSD(p, q) = H((p + q)/2) - (H(p) + H(q))/2$, where H is the Shannon entropy and zero shares contribute zero. The Bray– Curtis dissimilarity, equal to half the L1 distance for share vectors, was computed in parallel as a sensitivity metric. With n included countries a timepoint yields $n(n - 1)/2$ pairwise values, summarised by the mean, median and interquartile range.

Decomposition. Bray–Curtis dissimilarity was partitioned following Baselga into balanced variation in abundance and abundance gradients; for closed share vectors the gradient component is identically zero and the partition is reported as a mathematical identity. Shares were converted to presence (share ≥0.5%) and absence, and Jaccard dissimilarity was partitioned into turnover and nestedness components, with mean spectrum richness (pathogens present at ≥0.5% share) tracked per timepoint.

Per-pathogen dispersion and regional stratification. For each pathogen and timepoint the cross-country coefficient of variation and interquartile range of share were computed. Countries were mapped to the seven GBD super-regions using the standard GBD location hierarchy, and mean pairwise JSD was computed within each super-region (where at least five countries passed the filter) and between country pairs spanning different super-regions.

Statistics and software. Temporal trend was assessed by ordinary least squares regression of mean pairwise distance on calendar year across the five timepoints (three residual degrees of freedom), reporting the slope, Pearson correlation and two-sided p value, with two-sided alpha 0.05; the trend tests are interpreted as descriptive evidence of direction, and no adjustment was made for multiple correlated comparisons. Analyses used Python 3 with pandas, NumPy and SciPy.

Reproducibility. The revision analyses (joinpoint grid search, presence-threshold sensitivity grid, and the bootstrap of the 1990 to 2023 JSD change) are reproduced by a single released script (04_分析脚本/joinpoint_robustness_divergingspectra.py), with its full outputs provided as CSV files (Supplementary_Joinpoint_DivergingSpectra.csv, sensitivity_presence_threshold_DivergingSpectra.csv, bootstrap_jsd_change_DivergingSpectra.csv). The panel-construction and metric-decomposition scripts of the base analysis are available from the corresponding author on reasonable request.

decomposition metrics (run_metrics.py, run_metrics2.py), and figure generation (run_figures.py). Derived national share panels, pairwise distance series and analysis code are available from the corresponding author on reasonable request.
